# HIGH PRECISION CHARACTERIZATION OF RCCX REARRANGEMENTS IN A 21-HYDROXYLASE DEFICIENCY LATIN AMERICAN COHORT USING OXFORD NANOPORE LONG READ SEQUENCING

**DOI:** 10.1101/2024.11.14.24317161

**Authors:** Aldana Claps, Jorge Emilio Kolomenski, Franco Fernández, Natalia Macchiaroli, Marina L Ingravidi, Marisol Delea, Cecilia Fernández, Tania Castro, Julieta Laiseca, Laura Kamenetzky, Melisa Taboas, Liliana Dain

**Affiliations:** Centro Nacional de Genética Médica, Administración Nacional de Laboratorios e Institutos de Salud (ANLIS) “Dr. Carlos G Malbrán”, Buenos Aires, Argentina; Instituto de Biociencias, Biotecnología y Biología Traslacional (iB3), Facultad de Ciencias Exactas y Naturales, Universidad de Buenos Aires. Buenos Aires, Argentina; Instituto de Patología Vegetal (IPAVE-CIAP-INTA). Córdoba, Argentina; Unidad de Conocimiento Traslacional Hospitalaria Patagónica, Hospital de Alta Complejidad El Calafate SAMIC. Santa Cruz, Argentina; Laboratorio Novagen. Buenos Aires, Argentina

## Abstract

The gene *CYP21A2,* mapped to the RCCX module in 6p21.3, is responsible for the 21-hydroxylase deficiency (21OHD). In this work, we used Oxford Nanopore Technology (ONT) Long Reads (LR) sequencing to analyze samples from a large Argentinian cohort of 21OHD. Our goal was to gain additional information about the GVs involved in the rearrangements, to obtain higher resolution for the breakpoints of converted alleles and to retrieve new data of *TNX* and *CYP21A1P* genes. A total of 34 samples were sequenced in 2 amplicons of 8.5 Kb covering the centromeric and telomeric RCCX modules. The number of GVs found varied between 17-106 and all expected pathogenic GVs and new ones were obtained with the LR sequencing workflow developed. From the 21 alleles with macroconverted or chimeric genes containing the promoter of the *CYP21A1P*, we uncovered 5 of them lacking GVs commonly expected to be involved in 21OHD rearrangements. Importantly, we reclassified one patient with a deletion that had been overlooked with the current diagnostic technologies applied. These new findings change the genetic counseling for the patient and his family related to the presence of a carrier allele for Congenital Adrenal Hyperplasia and Ehlers-Danlos Syndrome (CAH-X). Furthermore, by addressing the study of the centromeric and the telomeric RCCX modules, we found 19 GVs for *CYP21A1P* and 29 GVs for *TNXA* not previously described in Latin American populations or not reported in any of the consulted databases. This study may represent the first one applying ONT LR in clinical studies in Latin America, highlighting the importance of LR sequencing as a high-resolution method of diagnosis and better cost-effective balance. It allows us to better understand the diversity of the RCCX modules and to gain knowledge of the molecular pathogenesis of the disease.

## INTRODUCTION

Congenital adrenal hyperplasia (CAH) due to 21-hydroxylase deficiency (21OHD), accounts for 90–95% of CAH cases. This autosomal recessive disorder, the most frequent inborn error of metabolism, has a broad spectrum of clinical forms, ranging from severe or classical (CL), including the salt-wasting (SW) and simple virilizing (SV) forms, to a mild late onset form or nonclassical (NC) [1].

The gene encoding 21-hydroxylase (21OHlasa), *CYP21A2,* is mapped to the short arm of chromosome 6 (6p21.3), within the human leukocyte antigen complex, with neighboring genes for tenascin (TNX), complement (C4), and the serine-threonine nuclear protein kinase 19 (formerly RP1). These four genes form a genetic unit with a length of 30 kb known as RCCX (*STK19 (RP1)-C4B-CYP21A2-TNXB*) module (Figure 1). Nearly two thirds of the chromosomes analyzed have a duplicated telomeric RCCX module which includes a genomic DNA segment composed of the pseudogenes *STK19B (RP2)-CYP21A1P-TNXA*, and another active copy of the C4A gene. *CYP21A1P* shares 98% sequence identity with *CYP21A2* [2–4]. Due to their high degree of sequence identity, unequal crossover during meiosis may generate large structural rearrangements and copy number changes in the RCCX module, containing both deletion and/or duplication of the gene *CYP21A2* and the pseudogene *CYP21A1*, chimeric genes *CYP21A1/CYP21A2* or *TNXB/TNXA*. To date, 11 different different chimeric *CYP21A1P*/*CYP21A2* genes have been described, being chimeric genes 1 to 9 the most frequently observed [5–7], In addition, 5 types of chimeras *TNXA*/*TNXB* have been described [8,9]. The chimeras *TNXA/TNXB* are associated with Congenital Adrenal Hyperplasia and Ehlers-Danlos Syndrome (CAH-X) that present joint hypermobility and a spectrum of other comorbidities associated with their connective tissue disorder, including chronic arthralgia, joint subluxations, hernias and cardiac defects [10]. In addition, gene conversion events may be responsible for the non-homologous transfer of a stretch of genetic variants from the pseudogene to the active gene that may result in alleles having micro or macroconversions based on the length of the conversion. Although most of the patients had pseudogene-derived pathogenic variants, an increasing number of novel and rare allelic variants have been found in disease-causing 21OHD alleles [1,11].

**Figure 1:**
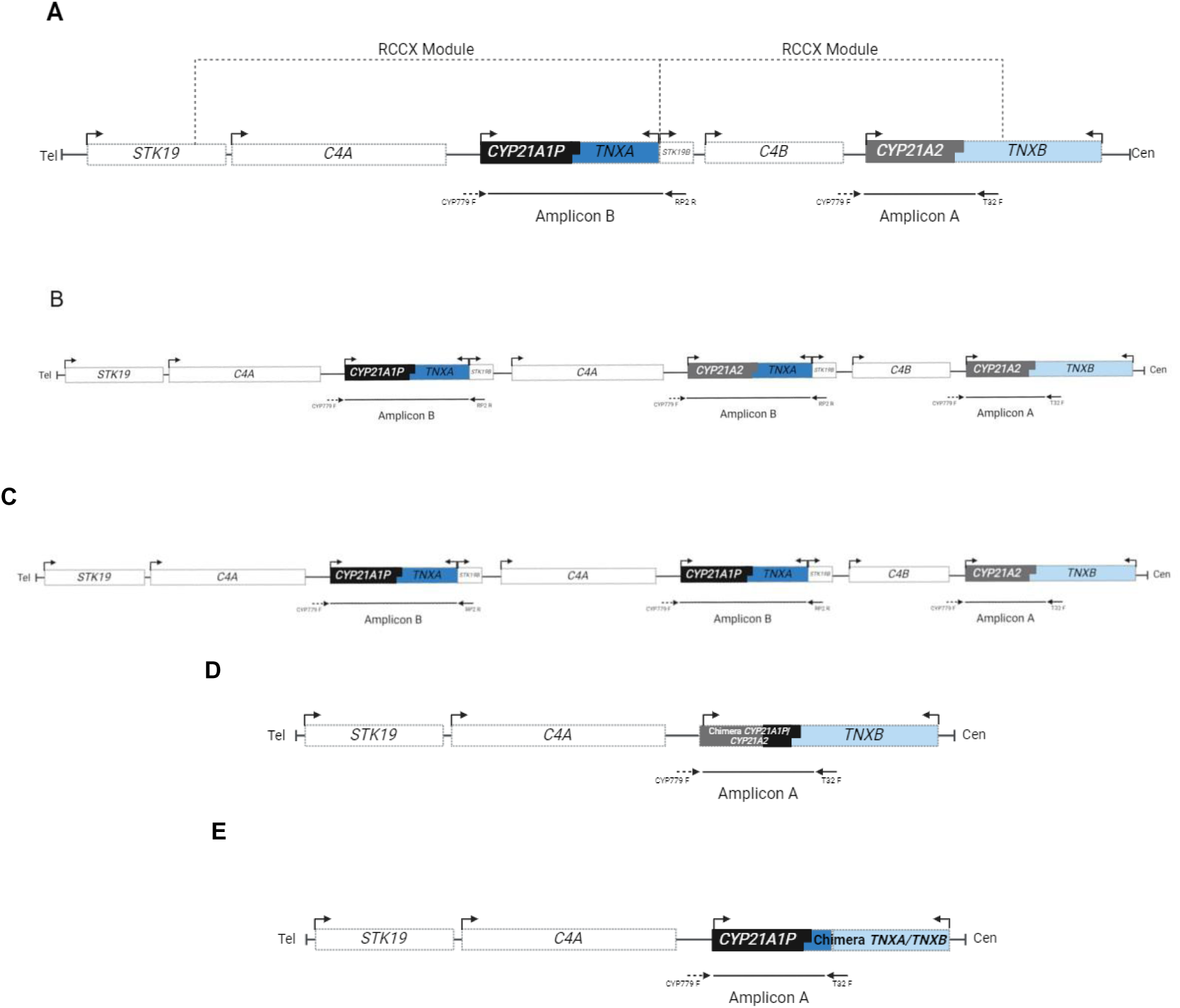
Structure of different arrangements of the RCCX modules. **A.** Bimodular rearrangement. **B**: Trimodular rearrangement containing a duplication of *CYP21A2*. **C**: Trimodular rearrangement containing a duplication of *CYP21A1P*. **D** and **E**: Structure of the RCCX module when the module containing *CYP21A2* is deleted. In D the product of the unequal recombination is a chimera *CYP21A1P*/*CYP21A2*, and in E, a chimera *TNXA*/*TNXB*. Boxes represent the genes: *TNX*: Tenascin X (B active gene, A: pseudogene). *C4 A* and *B*: genes of the complement factor 4. *CYP21A2* and *CYP21A1P*: gene and pseudogene, respectively: *STK19* and *STK19B*: active gene and pseudogene, respectively. Arrows above the boxes indicate orientation of transcription. Arrows below the boxes indicate the annealing position of the primers CYP779F, RP2R and Tena32F, to generate the 8,5 Kb amplicon A and amplicon B.

In a previous work, we performed a genetic characterization of a large cohort of 21OHD patients from Argentina. We used differential amplification of the *CYP21A2* gene and Sanger sequencing in 4 overlapping fragments together with Multiplex Ligation-dependent Probe Amplification (MLPA) to elucidate presence of pathogenic variants, as well as some of the possible genetic rearrangements in the RCCX region [12]. Nevertheless, these approaches are very time-consuming and not cost-effective, and do not reveal all the possible rearrangements of this complex genomic locus. They also do not include the search for putative pathogenic variants in the *TNXB* gene.

In this work we aim to leverage long read (LR) third-generation sequencing using Oxford Nanopore Technology (ONT) to analyze genetic variants (GVs) and arrangements in the RCCX region. LR has taken a more prominent role in recent years, especially by allowing for better analysis of genome rearrangements, repetitive sequences, regions with high sequence identity such as discriminating genuine genes from pseudogenes as well as to better elucidate *cis*/*trans* location of GVs [13]. Other methods based on short-read sequencing do not assemble it efficiently, so sequencing these regions is difficult to study [14]. We selected a group of samples previously studied in our laboratory as well as new samples recruited for this work, and sequenced both the centromeric and the telomeric RCCX modules. Our goal was to gain additional information about the GVs involved in the rearrangements, to obtain higher resolution for the breakpoints of converted and chimeric alleles and to retrieve new data of *TNXA* and *B* as well as *CYP21A1P* not previously studied.

## MATERIALS AND METHODS

### Ethical Approval

All the procedures performed in this study were in accordance with the 1964 Helsinki declaration and its later amendments or comparable ethical standards. Written informed consent was obtained from all individuals recruited for this study. The protocol was approved by the ethics committee of the Administración Nacional de Laboratorios e Institutos de Salud (A.N.L.I.S.), Buenos Aires, Argentina.

### Subjects

We selected a total of 34 DNA samples from our cohort for the study (Table 1). Twenty-eight were previously analyzed following the methodology established in our laboratory (samples 1-28) [12]. The selection of retrospective samples was based on their diversity in the RCCX module rearrangements and pathogenic GVs and compromised 12 samples from CL patients (3 SV, 9 SW), 11 NC, and 5 samples of relatives/partners/controls. In addition, we include 6 new DNA samples (samples 29-34) from 21OHlasa patients (1 SV and 5 NC) that were analyzed double blind using ONT LR and our current algorithm in parallel.

**Table 1:** Summary of the results obtained by LR sequencing in the 34 selected samples and its comparison using Sanger and MLPA.

| Sample ID | Condition | Presuntive phenotype | PREVIOUS RESULTS |  |  | LR RESULTS |  |  |  |  |  |
| --- | --- | --- | --- | --- | --- | --- | --- | --- | --- | --- | --- |
|  |  |  | Pathogenic GVs by Sanger sequencing | MLPA | 21A2 deduced genotype | AMPLICON A |  | AMPLICON B |  | Number of modules or alleles | Deduced genotype |
|  |  |  |  |  |  | Pathogenic GVs |  | Absence of frequent differentially pesudogen GVs / Duplicated 21A2 pathogenic GVs |  |  |  |
|  |  |  |  |  |  | HA1 | HA2 | HB1 | HB2 |  |  |
| 1 | Control | N/A | None | B / B | [WT];[WT] | None | None | 21A1P: p.V282L; p.R357W.<br>TNXA: c.12180C>G | - | B/M (21A1P del) or B/B' | [WT];[WT] |
| 2 | Patient | NC | p.V282L | B/ B<br>(21A1P/A2 Macroconv) | [p.V281L];<br>[Macroconv] | 21A2: p.V282L | 21A2: c.-126C>T; c.-113G>A; c.-110T>C; c.-103A>G; p.P31L; c.293-13A>G; p.G111Vfs*21; p.I173N; p.I237N-p.V238E-p.M240K; p.L308Ffs*6 | CYP21A1P: p.Q319*; p.R357W<br>TNXA: c.12530G>A; c.12520G>A; c.12180C>G | 21A1P: p.V282L; p.R357W | B/B | [p.V281L];[Macroconv] (equivalent to type 5) |
| 3 | Patient | NC | c.293-13A>AG; p.G111Vfs*21; p.V282L | T (21A1P dup)/B (21A1P/A2 5'Macroconv) or B/T (21A1P/A2 5'Macroconv; 21A1P dup) | [p.V281L];<br>[5'Macroconv] | 21A2: p.V282L | 21A2: c.-126C>T; c.-113G>A; c.-110T>C; c.-103A>G; c.-4C>T; p.P31L; c.293-13A>G; p.G111Vfs*21 | TNXA: c.12530G>A² | 21A1P: p.V282L; p.Q319*; p.R357W.<br>TNXA: c.12180C>G | B/T (21A1P dup) | [p.V281L];[5'Macroconv] (equivalent to type 1) |
| 4 | Patient | CL (SW) | c.293-13C>G; p.G111Vfs*21 | T (21A1P/A2 5'Macroconv; 21A1P dup)/M (Ch TNXA/B type 1) or B (21A1P/A2 5'Macroconv)/ B (Macroconv TNXA/B type 1) | [5'Macroconv];[Ch or Macroconv TNXA/B type 1] | 21A2: c.-126C>T; c.-113G>A; c.-110T>C; c.-103A>G; c.-4C>T; p.P31L; c.293-13C>G; p.G111Vfs*21 | 21A2: c.-126C>T; c.-113G>A; c.-110T>C; c.-103A>G; c.-4C>T; p.P31L; c.293-13C>G; p.G111Vfs*21; p.I173N; p.I237N-p.V238E-p.M240K; p.V282L; p.L308Ffs*6; p.R357W<br>TNXB: c.12530G>A; c.12224G>A; c.12180C>G c.11435_11524+30del | 21A1P: p.Q319*<br>TNXA: c.12520G>A | None | T (21A1P dup)/M (Ch TNXA/B ) or B/B | [5'Macroconv](equivalent to type 1);[Ch or Macroconv TNXA/B type 1] |
| 5 | Patient | CL (SW) | c.293-13C>G; p.G111Vfs*21 | M/M (Ch 21A1P/A2 type 1/ Ch 21A1P/A2 type 1) | [Ch 21A1P/A2 type 1];[Ch 21A1P/A2 type 1] | 21A2: c.-126C>T; c.-113G>A; c.-110T>C; c.-103A>G; c.-4C>T; p.P31L; c.293-13C>G; p.G111Vfs*21 | 21A2: c.-126C>T; c.-113G>A; c.-110T>C; c.-103A>G; c.-4C>T; p.P31L; c.293-13C>G; p.G111Vfs*21 | No amplification |  | M/M | [Ch 21A1P/A2 type 1];[Ch 21A1P/A2 type 1] |
| 6 | Partner | N/A | None | B/M (21A1P del) | [WT];[WT] | None | None | None | - | B/M (21A1P del) or B/B' | [WT];[WT] |
| 7 | Partner | N/A | c.*13G>A; p.Q319* | M (21A1P del)/ T (21A2 dup) | [c.*13G>A ];[ p.Q319*;WT] | 21A2: c.*13G>A | 21A2: p.Q319*<br>TNXB: c.12469+2T>C | 21A1P: p.P31L; p.I173N; p.R357W<br>TNXA: c.12224G>A | 21A2 WT | T (21A2 dup)/ M (21A1P del) or T (21A2 dup)/ B¹ | [c.*13G>A];[p.Q319*;c.12469+2T>C;WT] |
| 8 | Patient | CL (SW) | p.G111Vfs*21 | M (Ch 21A1P/A2 type 5) / B (p.G111Vfs*21) | [Ch 21A1P/A2 type 5];[p.G111Vfs*21] | 21A2: c.-126C>T; c.-113G>A; c.-110T>C; c.-103A>G; p.P31L; c.293-13C>G; p.G111Vfs*21; p.I173N; p.I237N-p.V238E-p.M240K; p.L308Ffs*6 | 21A2: p.G111Vfs*21 | None | - | M (Ch 21A1P/A2) / B or B (Macroconv)/B¹ | [Ch or macroconv 21A1P/A2 type 5];[p.G111Vfs*21] |

| Sample ID | Condition | Presuntive phenotype | PREVIOUS RESULTS |  |  | LR RESULTS |  |  |  |  |  |
| --- | --- | --- | --- | --- | --- | --- | --- | --- | --- | --- | --- |
|  |  |  | Pathogenic GVs by Sanger sequencing | MLPA | 21A2 deduced genotype | AMPLICON A |  | AMPLICON B |  | Number of modules or alleles | Deduced genotype |
|  |  |  |  |  |  | Pathogenic GVs |  | Absence of frequent differentially pesudogen GVs / Duplicated 21A2 pathogenic GVs |  |  |  |
|  |  |  |  |  |  | HA1 | HA2 | HB1 | HB2 |  |  |
| 9 | Patient | CL (SW) | c.293-13C>G; p. G111Vfs*21 | B/ T (21A1P dup) | [c.293-13C>G]; [5'Macroconv] | 21A2:<br>c.293-13C>G | 21A2:<br>c.-126C>T; c.-113G>A; c. -110T>C; c.-103A>G; c.-4 C>T; p.P31L; c.293-13C>G; p.G111Vfs*21 | None<sup>2</sup> | 21A1P:<br>p.V282L; p.R357W<br>TNXA:<br>c.12180C>G | B/T (21A1P dup) | [c.293-13C>G]; [5'Macroconv] (equivalent to type 1) |
| 10 | Patient | CL (SW) | p.L308Ffs*6 | M (Ch 21A1/A2 type 5) / B (p.L308Ffs*6) | [Ch 21A1P/A2 type 5];[p.L308Ffs*6] | 21A2: c.-126C>T; c.-113 G>A; c.-110T>C; c.-103 A>G;c.-4C>T; p.P31L; c. 293-13C>G; p. G111Vfs*21; p.l173N; p. l237N-p.V238E-p.M240K; p.V282L; p.L308Ffs*6; p. Q319*; p.R357W<br>TNXB: c.12520G>A; c. 12224G>A; c.12180C>G | 21A2:<br>p.L308Ffs*6 | TNXA:<br>c.12530G>A; c. 12520G>A; c. 12224G>A | - | M (Ch TNXA/B)/B or B (Macroconv)/B<sup>1</sup> | [Ch or Macroconv TNXA- TNXB type 2 ];[p. L308Ffs*6] |
| 11 | Patient | NC | None | B (21A1P/A2 Macroconv)/B or M (Ch 21A1P/A2 type 5) / T (21A1P dup) | [Macroconv];[WT] or [Ch 21A1P/A2 type 5];[WT] | CYP21A2: c.-126C>T; c. -113G>A; c.-110T>C; c. -103A>G;c.-4C>T; p.P31L; p.H63L; c.293-13C>G; p. G111Vfs*21; p.l173N; p. l237N-p.V238E-p.M240K; p.V282L; p.L308Ffs*6; p. Q319* | None | None | 21A1P<sup>3</sup>:<br>p.R357W | B / B or T (21A1P dup) / M (Ch 21A1P/A2) | [Ch or Macroconv A1P/A2 type 3];[WT] |
| 12 | Patient | NC | p.V282L | T (21A1P dup)/ B | [p.V282L];[p.V282L] | 21A2:<br>p.V282L | - | 21A1P:<br>p.V282L; p.R357W | 21A1P:<br>p.Q319*<br>TNXA:<br>c.12530G>A; c. 12520G>A; c. 12180C>G | B / B | [p.V282L];[p.V282L] |
| 13 | Patient | CL (SW) | c.293-13C>G; p. G111Vfs*21 | M (Ch 21A1P-21A2 type 1) / M (Ch 21A1P-21A2 type 1) | [Ch 21A1P/A2 type 1];[Ch 21A1P/A2 type 1] | 21A2: c.-126C>T; c.-113 G>A; c.-110T>C; c.-103 A>G; c.-4C>T; p.P31L; c. 293-13C>G; p. G111Vfs*21 | 21A2: c.-126C>T; c.-113G>A; c.-110T>C; c.-103A>G; c.-4 C>T; p.P31L; c.293-13C>G; p. G111Vfs*21 | No amplification |  | M (Ch21A1P/A2) / M (Ch 21A1P/A2) | [Ch 21A1P/A2 type 1];[Ch 21A1P/A2 type 1] |
| 14 | Patient | CL (SW) | c.293-13C>G; p. G111Vfs*21; p.R445* | B / M (Ch 21A1P- 21A2 type 1) | [p.R445*];[Ch 21A1P/A2 type 1] | 21A2:<br>p.R445* | CYP21A2: c.-126C>T; c.-113 G>A; c.-110T>C; c.-103A>G; c.-4C>T; p.P31L; c.293- 13C>G; p.G111Vfs*21 | 21A1P:<br>p.V282L; p.R357W | - | M (Ch 21A1P/A2)/B or B/B<sup>1</sup> | [p.R445*];[Ch or Macroconv 21A1P/A2 type 1] |
| 15 | Patient | NC | c.-126C>T;c.-113G>A; c.-110T>C; c.-103 A>G; c.-4C>T; p.P31L | B/B (21A2 5' Macroconv type 4 or 9) | [p.P31L]; [5'Macroconv (equivalent to type 4)] | 21A2: c.-126C>T; c.-113 G>A; c.-110T>C; c.-103 A>G; c.-4C>T; p.P31L | 21A2:<br>p.P31L | None | TNXA:<br>c.12530G>A; c. 12520G>A; c. 12224G>A | B/ B | [5'Macroconv](equivalent to type 4);[p.P31L] |
| 16 | Patient | NC | p.V282L | T (21A1P dup) / T (21A1P dup) | [p.V282L];[p.V282L] | CYP21A2:<br>p.V282L | - | 21A1P:<br>p.Q319*; p.R357W<br>TNXA:<br>c.12530G>A; c. 12520G>A; c. 12180C>G | CYP21A1P:<br>p.V282L; p.R357W | B/B | [p.V282L];[p.V282L] |

| Sample ID | Condition | Presuntive phenotype | PREVIOUS RESULTS |  |  | LR RESULTS |  |  |  |  |  |
| --- | --- | --- | --- | --- | --- | --- | --- | --- | --- | --- | --- |
|  |  |  | Pathogenic GVs by Sanger sequencing | MLPA | 21A2 deduced genotype | AMPLICON A |  | AMPLICON B |  | Number of modules or alleles | Deduced genotype |
|  |  |  |  |  |  | Pathogenic GVs |  | Absence of frequent differentially pesudogen GVs / Duplicated 21A2 pathogenic GVs |  |  |  |
|  |  |  |  |  |  | HA1 | HA2 | HB1 | HB2 |  |  |
| 17 | Patient | NC | p.Q319* | T (21A2 dup)/M (21A1P del) | [p.Q319*;WT];[WT] | None | 21A2:<br>p.Q319*<br>TNXB:<br>c.12469+2T>C | 21A1P: p.P31L; p. I173N; p.R357W<br>TNXA:<br>c.12224G>A | 21A2 WT | T(21A2 dup) / M (21A1P del) or<br>T (21A2 dup) / B¹ | [p.Q319* ; c.12469+2T>C; WT];[WT] |
| 18 | Patient | CL (SW) | c.293-13C>G; p. G111Vfs*21 | B (c.293-13C>G) / B (21A2 5' Macroconv) | [c.293-13C>G]; [5' Macroconv] | CYP21A2:<br>c.293-13C>G | CYP21A2:<br>c.-126C>T; c.-113G>A; c. -110T>C; c.-103A>G; c.-4 C>T; p.P31L; c.293-13C>G; p.G111Vfs*21 | None | - | B / B¹ or<br>B / M (Ch 21A1P/A2) | [c.293-13C>G]; [5' Macroconv] (equivalent to type 1) or [c.293-13C>G];[Ch 21A1P/A2 type 1] |
| 19 | Patient | CL (SV) | c.293-13C>G; p. V282L; p.L308Ffs*6; p.Q319* ; p.R357W | B (c.293-13C>G) / B (21A2 3' Macroconv) | [c.293-13C>G]; [3' Macroconv] | 21A2:<br>c.293-13C>G | 21A2:<br>p.V282L; p.L308Ffs*6; p. Q319* ; p.R357W | 21A1P*:<br>c.-4C>T; p.P31L<br>TNXA:<br>c.12530G>A | 21A1P: p.R357W<br>TNXA: c. 12530G>A; c. 12180C>G | B / B | [c.293-13C>G]; [3' Macroconv] |
| 20 | Patient | NC | p.I237N-p.V238E-p. M240K; p.V282L; p. L308Ffs*6 | T (dup 21A1P)/ B (21A2 3' Macroconv) | [p.V282L]; [3' Macroconv] | CYP21A2:<br>p.V282L | 21A2: p.I237N-p.V238E-p. M240K; p.V282L; p. L308Ffs*6 | 21A1P: p.Q319* ; p.R357W | 21A1P²: p.V282L; p.R357W<br>TNXA:<br>c.12180C>G | T (21A1P dup)/ B | [p.V282L];[3' Macroconv] |
| 21 | Patient | NC | p.V282L | B/ B (21A1P/A2 Macroconv equivalent to type 5) | [p.V282L]; [Macroconv] | CYP21A2:<br>p.V282L | 21A2:<br>c.-126C>T; c.-113G>A; c. -110T>C; c.-103A>G; p.P31L; c.293-13A>G; p.G111Vfs*21; p.I173N; p.I237N-p.V238E-p. M240K; p.L308Ffs*6 | 21A1P: p.Q319* ; p.R357W<br>TNXA:<br>c.12530G>A | 21A1P: p.V282L; p.R357W | B/B | [p.V282L];[Macroconv] (equivalent to type 5) |
| 22 | Patient | CL (SV) | p.I173N | M (Ch TNXA/B type 1)/B | [p.I173N];[Ch TNXA/B type 1] | 21A2:<br>p.I173N | 21A2: c.-126C>T; c.-113G>A; c.-110T>C; c.-103A>G; c.-4 C>T; p.P31L; c.293-13A>G; p.G111Vfs*21; p.I173N; p. I237N-p.V238E-p.M240K; p. V282L; p.L308Ffs*6; p. Q319* ; p.R357W<br>TNXB: c.12530G>A; c. 12520G>A; c.12224G>A; c. 12180C>G; c. 11435_11524+30del | None | - | B /M (Ch TNXA/B) or<br>B/B¹ | [p.I173N];[Ch or Macroconv TNXA/B type 1] |
| 23 | Relative | N/A | c.293-13C>G; p. G111Vfs*21; p.Q319* | T (21A2 dup) / B 5' Macroconv equivalent to type 1) | [p.Q319* ; WT]; [5' Macroconv] (equivalent to type 1) | 21A2:<br>p.Q319*<br>TNXB:<br>c.12469+2T>C | 21A2:<br>c.-126C>T; c.-113G>A; c. -110T>C; c.-103A>G; c.-4 C>T; p.P31L; c.293-13C>G; p.G111Vfs*21 | None | 21A2 WT | B/B | [p.Q319* ; c.12469+2T>C; WT];[5' Macroconv] (equivalent to type 1) or [p.Q319* ; c.12469+2T>C; WT];[Ch 21A1P/2 type 1] |
| 24 | Patient | CL (SV) | c.293-13C>G; p. H403Rfs*5 | B/ M(21A1P del) | [c.293-13C>G];[p. H403Rfs*5] | 21A2:<br>c.293-13C>G | 21A2:<br>p.H403Rfs*5 | None | - | B/ M (21A1P del) or B¹ | [c.293-13C>G];[p. H403Rfs*5] |
| 25 | Relative | N/A | c.293-13C>G; p. Q319* | T(21A2 dup) / B | [c.293-13C>G; p. Q319*]; [WT] | None | 21A2:<br>p.Q319*<br>TNXB:<br>c.12469+2T>C | 21A1P²: p.I173N; p.V282L; p.R357W<br>TNXA:<br>c.12180C>G | 21A2: c.293-13C>G | T (21A2 dup) / B | [p.Q319* ; c.12469+2T>C ; c.293-13C>G];[WT] |

| Sample ID | Condition | Presuntive phenotype | PREVIOUS RESULTS |  |  | LR RESULTS |  |  |  |  |  |
| --- | --- | --- | --- | --- | --- | --- | --- | --- | --- | --- | --- |
|  |  |  | Pathogenic GVs by Sanger sequencing | MLPA | 21A2 deduced genotype | AMPLICON A |  | AMPLICON B |  | Number of modules or alleles | Deduced genotype |
|  |  |  |  |  |  | Pathogenic GVs |  | Absence of frequent differentially pesudogen GVs / Duplicated 21A2 pathogenic GVs |  |  |  |
|  |  |  |  |  |  | HA1 | HA2 | HB1 | HB2 |  |  |
| 26 | Patient | CL (SW) | No amplification | B (Macroconv equivalent to type 5) / M (Ch 21A1P/A2 type 5) | [Macroconv (equivalent to type 5)]; [Ch 21A1P/A2 type 5] | 21A2:<br>c.-126C>T; c.-113G>A; c.-110T>C; c.-103A>G; c.-4C>T; p.P31L; p.H63L; c.293-13A>G; p.G111Vfs*21; p.I173N; p.I237N-p.V238E-p.M240K; p.V282L; p.L308Ffs*6; p.Q319* | - | 21A1P<sup>3</sup> :<br>p.R357W | - | B (Macroconv / M (Ch CYP21A1P/A2) or B (Macrocon)/B (Macroconv)<sup>1</sup> | [Macroconv];[Ch 21A1P/A2 type 3] |
| 27 | Patient | NC | p.I173N; p.I237N-p.V238E-p.M240K; p.V282L; p.L308Ffs*6; p.Q319* | B / B (Macroconv TNXA/B type1) | [WT]; [Macroconv TNXA/B type1] | CYP21A2: c.-126C>T; c.-113G>A; c.-110T>C; c.-103A>G; c.-4C>T; p.P31L; c.293-13C>G; p.I173N; p.I237N-p.V238E-p.M240K; p.V282L; p.L308Ffs*6; p.Q319*<br>TNXB: c.12530G>A; c.12224G>A; c.12180C>G; c.11435_11524+30del | None | 21A1P:<br>p.G111fs; p.R357W<br>TNXA:<br>c.12180C>G | None | B / B (macronv TNXA/B) or B/Ch TNXA/B | [WT];[Macroconv or Ch TNXA/B] |
| 28 | Patient | NC | p.V282L | T (21A1P dup) / T (21A1P dup) | [p.V282L];[p.V282L] | CYP21A2:<br>p.V282L | - | 21A1P:<br>p.Q319*; p.R357W<br>TNXA:<br>c.12530G>A; c.12520G>A; c.12180C>G | CYP21A1P:<br>p.V282L; p.R357W | B / B | [p.V282L];[p.V282L] |
| 29 | Patient | NC | p.R484P; p.V282L | B/ T (21A1P dup) | [p.R484P];[p.V282L] | CYP21A2:<br>p.R484P | CYP21A2:<br>p.V282L | None | CYP21A1P:<br>p.V282L; p.Q319*; p.R357W<br>TNXA:<br>c.12180C>G | Bimodular / Trimodular (CYP21A1P duplication) | [p.R484P];[p.V282L] |
| 30 | Patient | NC | p.I237N-p.V238E-p.M240K; p.V282L; p.L308Ffs*6 | B/ T (21A1P dup) | [3' Macroconv];[p.V282L] | 21A2:<br>p.I237N-p.V238E-p.M240K; p.V282L; p.L308Ffs*6 | 21A2:<br>p.V282L | 21A1P<sup>2</sup>:<br>p.Q319; p.R357W | 21A1P:<br>p.V282L; p.Q319; p.R357W<br>TNXA:<br>c.12180C>G | B / T (21A1P dup)<sup>2</sup> | [3' Macroconv];[p.V282L] |
| 31 | Patient | NC | p.V282L | T (21A1P dup) / T (21A1P dup) | [p.V282L];[p.V282L] | 21A2:<br>p.V282L | - | CYP21A1P: p.Q319*; p.R357W<br>TNXA:<br>c.12530G>A; c.12520G>A; c.12180C>G | 21A1P: p.V282L; p.R357W | B / B | [p.V282L];[p.V282L] |
| 32 | Patient | NC | p.V282L; p.I173N | B / T (21A1P dup) | [p.I173N];[p.V282L] | 21A2:<br>p.I173N | 21A2:<br>p.V282L | None<sup>2</sup> | 21A1P: p.Q319*; p.R357W<br>TNXA: c.12530G>A; c.12520G>A; c.12180C>G | B / T (21A1P dup) | [p.I173N];[p.V282L] |

| Sample ID | Condition | Presuntive phenotype | PREVIOUS RESULTS |  |  | LR RESULTS |  |  |  |  |  |
| --- | --- | --- | --- | --- | --- | --- | --- | --- | --- | --- | --- |
|  |  |  | Pathogenic GVs by Sanger sequencing | MLPA | 21A2 deduced genotype | AMPLICON A |  | AMPLICON B |  | Number of modules or alleles | Deduced genotype |
|  |  |  |  |  |  | Pathogenic GVs |  | Absence of frequent differentially pesudogen GVs / Duplicated 21A2 pathogenic GVs |  |  |  |
|  |  |  |  |  |  | HA1 | HA2 | HB1 | HB2 |  |  |
| 33 | Patient | NC | p.V282L | T (21A1P dup) / T (21A1P dup) | [p.V282L];[p.V282L] | 21A2: p.V282L | - | 21A1P: p.Q319*; p.R357W<br>TNXA: c. 12530G>A; c. 12520G>A; c. 12180C>G | 21A1P: p.V282L; p.R357W | B/B | [p.V282L];[p.V282L] |
| 34 | Patient | CL (SV) | c.293-13C>G | B/B | [c.293-13C>G];[c.293-13C>G] | 21A2: c.293-13C>G | 21A2: c.293-13C>G | None | - | B / M (CYP21A1P deletion) or B/B¹ | [c.293-13C>G];[c.293-13C>G] |

CAH: Congenital adrenal hyperplasia. CL: Classical, SW: salt-wasting, SV: simple virilizing, NC: Nonclassical. GV's: Genetic Variants. LR: Long Reads. MLPA: Multiplex Ligation-dependent Probe Amplification. HA1 and HA2: Haplotype 1 and haplotype 2
for amplicon A; HB1 and HB2, haplotypes 1 and 2 for amplicon B. WT: Wild Type. B. Bimodular, M: Monomodular, T: Trimodular. del: deletion; dup: duplication. Macroconv: macroconversion. Ch: Chimera. N/A: Not Applicable. A dash (-) denotes that only one haplotype could be phased. 1: Homozygosity for *CYP21A1P*. 2: Heterozygous GV's in HB1. 3. *CYP21A1P* presented the p.63L GV. The GV's are referenced to *CYP21A2*: RefSeq for cDNA: NM\_000500.9; protein: NP\_000491.4; TNXB: RefSeq for cDNA: NM\_001365276.2 and Ensembl transcript: ENST00000644971.2; protein: NP\_001352205.1; CYP21A1P: Ensembl for non-coding transcript ENST00000354927; TNXA: Ensembl for non-coding transcript ENST00000507684.1.

### Long Range PCR

For each sample, 2 different fragments (Amplicon A and B) of 8,5 kb were amplified using primers CYP779F and TENA32F or CYP779F and RP2R [15,16] (Figure 1). Amplicon A includes the module containing *CYP21A2*-*TNXB* (exon 32-44 of *TNXB*) while amplicon B includes the module/s containing *CYP21A1P-TNXA-STK19B(RP2)* and/or *CYP21A2-TNXA-STK19B(RP2)*. Long range PCRs were performed using 3 μL of 300 ng of genomic DNA, 3.75 UI of Expand Long Polymerase (Expand Long Template PCR System Kit, RocheⓇ), 1.75 mM of Expand Long Template buffer 3 (Expand Long Template PCR System Kit, RocheⓇ), 350 μM of dNTPs, 300 nM of each primer and PCR grade water up to 50 μL. Cycling conditions were: 94 °C denaturation for 2 min followed by 10 cycles of 94 °C denaturation for 10 s, 60 °C annealing for 30 s, 68 °C elongation for 9 min, followed by 25 cycles of 94 °C denaturation for 15 s, 60 °C annealing for 30 s, 68 °C elongation for 9 min adding 20 s cycle elongation for each successive cycle and a final elongation of 7 min at 68 °C. All the PCR products were detected by electrophoresis on a 1.0% agarose gel and ethidium bromide stain.

### Long-read sequencing

After amplification, each amplicon was purified using 1X Agencourt AMPure XP beads (Beckman CoulterⓇ) and quantified using the Qubit dsDNA High Sensitivity Reagent (ThermoFisher ScientificⓇ) while quality was evaluate using NanoDrop™ (ThermoFisher ScientificⓇ). DNA repair and end-prep were performed using 200 fmol of each amplicon and NEBNext Ultra II End repair/dA-tailing Module (E7546-New England BiolabsⓇ), Native barcode ligation were performed using NEB Blunt/TA Ligase Master Mix (M0367-New England BiolabsⓇ).Library preparation, barcoding and sequencing were performed with Native barcoding amplicons kit (EXP-NBD104, EXP-NBD114, and SQK-LSK109) for MinION-based long-read sequencing using Nanopore Flow Cell R9.4.1 (number 1-24) and Native barcoding Kit 24 V14 (SQK-NBD114.24) for PromethION-based long-read sequencing using Nanopore Flow Cell R10.4.1 (number 25-34), following ONT recommendations. Amplicon A and B from each sample had a different barcode.

We developed a set of custom scripts in Python based on the recommended ONT pipelines for the analysis of LR. We used Guppy version 6.5.7 (Oxford Nanopore Technologies Ltd., 2000, [17] for basecalling and barcoding of the reads, Minimap2 version 2.17 [18], Miniasm version 0.3 [19] and Samtools version 1.3.1 [20]) to generate the BAM files with the reads aligned to reference sequences, and Clair3 version 1.0.3 [21] and Whatshap version 2.0 [22] for variant calling and haplotyping. The reads were aligned to both reference sequences of each amplicon and the human genome (hg38). When aligning to the GRCh38 human genome reference sequence (hg38), amplicon A corresponds to positions chr6:32037620 to chr6:32046165, whereas amplicon B corresponds to positions chr6:32004884 to chr6:32013454. Figure 2 summarizes the workflow applied for the LR sequencing and analysis.

**Figure 2:**
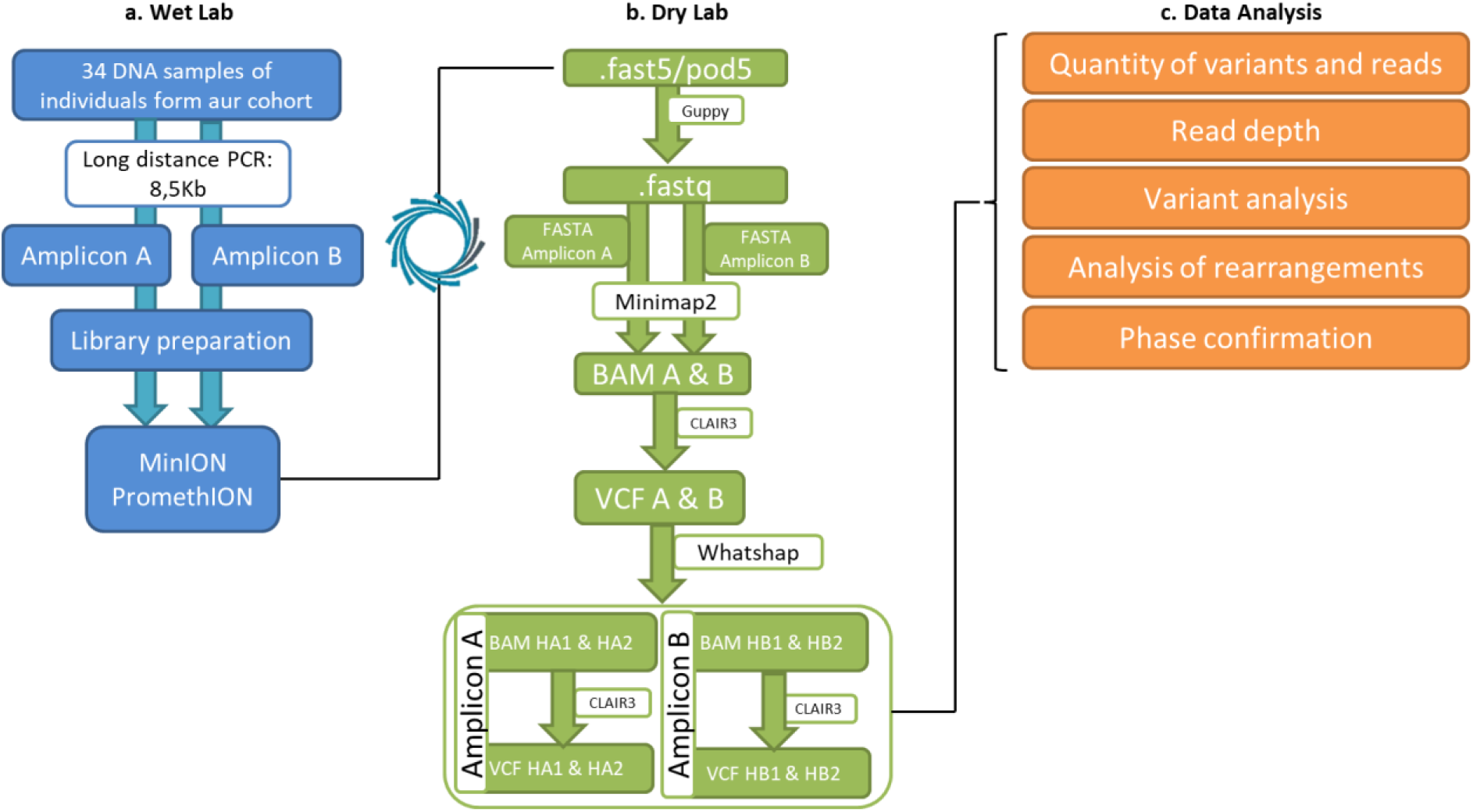
Summary of the workflow. (a) Wet Lab optimization. Both amplicons of the RCCX module were amplified by PCR and sequenced using a MinION or PromethION device. (b) Dry Lab optimization. Fast5 or pod5 files were processed using Guppy, Minimap2, Clair3 and Whatshap to obtain bam and vcf files. (c) Data analysis. Analysis of ONT sequencing output. HA1 and HA2: Haplotype 1 and haplotype 2 for amplicon A, respectively. HB1 and HB2: Haplotype 1 and haplotype 2 for amplicon B, respectively.

Nomenclature of the GVs were following the recommendations of the Human Genome Variation Society (HGVS) [23] according to NC_000006.12 reference sequence from GRCh38.p14 (*CYP21A2*: RefSeq for cDNA: NM_000500.9 and Ensembl transcript: ENST00000418967; protein: NP_000491.4; *TNXB*: RefSeq for cDNA: NM_001365276.2 and Ensembl transcript: ENST00000644971.2; protein: NP_001352205.1 ; *CYP21A1P*: Ensembl for non-coding transcript ENST00000354927; *TNXA*: Ensembl for non-coding transcript ENST00000507684.1; *STK19B*: GenBank assembly GCF_000001405.40). Chimeric genes were classified according to the different breakpoints involved in the deletion [5,9]. Large gene conversions were named as macroconversions; 5’ and 3’ macroconversions referred to alleles converted up to or 3’ from the p.G111Vfs*21 GV, respectively. The following population databases were consulted to retrieve information and frequencies of the different GVs: GnomAD (https://gnomad.broadinstitute.org/, version 2.1.1 [24], 1000 genome project (https://www.internationalgenome.org/, [25]), dbSNP (https://www.ncbi.nlm.nih.gov/snp/, [26] including dbGaP dataset derivate from Allele Frequency Aggregator (ALFA) project [27]. In addition, ClinVar database (https://www.ncbi.nlm.nih.gov/clinvar/, [28], ClinGen database (https://www.clinicalgenome.org/, [29], DECIPHER browser (https://www.deciphergenomics.org/, [30] and PDB database (Protein Data Bank: https://www.rcsb.org/) was also used for analysis of GVs. The novel variant found was deposited in ClinVar (submission ID: SUB14707012) and the classification was following the recommendations of the American College of Medical Genetics (ACMG) guideline [31].

## RESULTS

We used third-generation ONT LR sequencing to analyze 34 samples from an Argentinian cohort of 21-hydroxylase deficiency. Twenty-eight samples were previously studied in our lab by Sanger sequencing and MLPA and the remaining 6 were studied for the first time in this work. To this end, we amplified and sequenced 2 amplicons of 8,5 kb spanning the *CYP21A2-TNXB* genes (amplicon A) and *CYP21A1P-TNXA* pseudogenes or a duplicated *CYP21A2-TNXA* (amplicon B), respectively. For each amplicon, we analyzed the presence of pathogenic and non-pathogenic GVs, indels, and their zygosity after haplotyping and phasing, and defined large gene conversions or chimeric alleles.

We successfully sequenced both amplicons in 32/34 samples. The remaining 2 samples were genotyped as chimera/chimera, and as expected, only amplicon A showed amplification (see below). The read depth was >400X for all the amplicons, and the estimated N50 was 8,5kb for all sequence output.

The quality cutoff of reads was >Q5 for samples 1-24 and >Q7 for samples 25-34. In addition, we performed a comparison of the average basecalling quality score of the amplicon A of 4 samples genotyped as p.V282L/p.V282L (Supplementary Figure S1). Samples 12 and 16 were sequenced by a MinION device using Nanopore Flow Cell R9.4.1 while samples 28 and 31 were sequenced by a PromethION device with Nanopore Flow Cell R10.4.1. Median read quality for R9 chemistry was 12,25 and 16,55 for R10.

For Minion based sequencing, the number of GVs found varied between 17-106 in amplicon A, and between 3-66 for amplicon B, while for PromethION based sequencing the number of GVs ranged between 12-80 and 15-55 for amplicon A and B, respectively. Excluding large rearrangements, we found a total of 248 single nucleotide variants (SNVs), 3 insertions, 6 deletions and 11 duplications (Supplementary Table S1 and S2). Particularly, for the coding regions of *CYP21A2* and *TNXB*, we retrieved 17 synonymous SNVs (9 in *CYP21A2* and 8 in *TNXB),* 28 missense (15 in *CYP21A2* and 13 in *TNXB*), 5 frameshift (4 in *CYP21A2* and 1 in *TNXB*), 1 inframe and 2 nonsense in *CYP21A2.* In addition, we found 2 duplications and 3 deletions in *CYP21A2* and 1 duplication and 1 deletion in *TNXB*. Finally, we observed 39 SNVs in *CYP21A1P* and 54 in *TNXA*, and 2 duplications, 1 insertion and 1 deletion in *CYP21A1P*.

The results of genotyping by ONT LR sequencing are summarized in Table 1, along with the results obtained applying our current diagnosis methodology. As shown, all the pathogenic GVs found by Sanger sequencing in *CYP21A2* were indeed observed after ONT LR sequencing in all the analyzed samples. Common non-pathogenic variants (synonymous, non-synonymous, indels) were also concordant using both approaches (data not shown). LR sequencing, however, allowed us to pricisely phase cis/trans locations of GVs (see Figure 3 for some examples), and to complete the list of GVs involved in the rearrangements that were not detected previously with the methodology used (Table 1).

**Figure 3:**
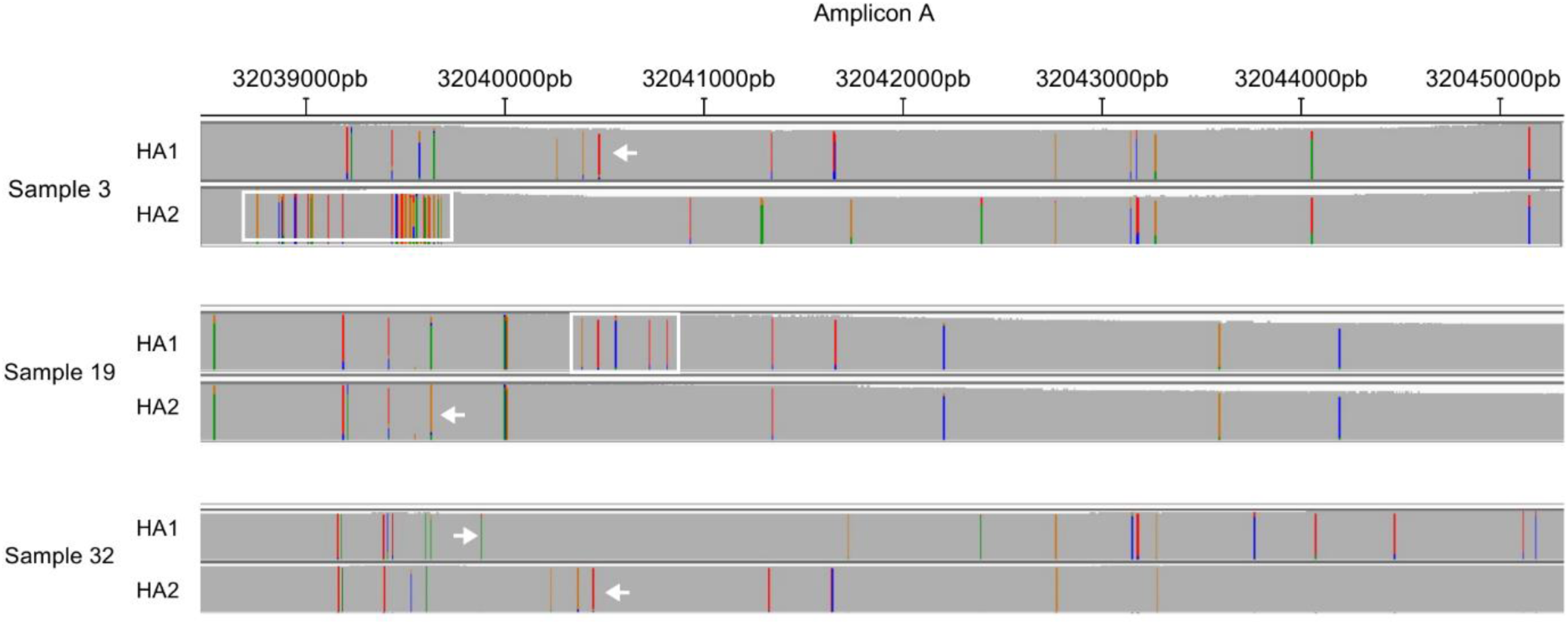
Representative IGV plots showing cis/trans location of variants of samples ID 3, 19 and 32 for amplicon. **A**. The arrows indicate the GVs p.V282L for haplotype 1 (HA1) in sample 3, c.293-13C>G for haplotype 2 (HA2) in sample 19 and p.I173N and p.V282L for HA1 in sample 32. The rectangle indicates the 5’ macroconversion in HA2 for sample 3 and the 3’ macroconversion in HA1 for sample 19. Above the plot are the genomic locations in hg38 reference sequence.

After analyzing the results, we found that some of the common *CYP21A1P* pseudogene GVs were absent in the rearrangements. Among the 21 alleles that had a macroconverted or chimeric gene that included the promoter of the *CYP21A1P* and different lengths of the coding region, 3 alleles with a breakpoint at least at the p.L308Ffs*6, lacked the c.-4C>T variant at the promoter and the p.V282L GV (samples 2, 8 and 21). In 1 allele (sample 4), the chimera reaches the *TNXB* gene, although the p.Q319* variant was not observed. Likewise, we observed a *TNXA/TNXB* converted allele lacking the p.G111Vfs*21 GV as does the pseudogene of the same sample (sample 27). Similarly, some common GVs in *TNXA*, are absent in the converted or chimeric alleles (Table 1).

In addition, by analyzing the entire region in a single read, the genomic region involved in the rearrangements could be more accurately defined (Figure 4). In that sense, the start point of the breakpoint for type 1 *CYP21A1P*/*CYP21A2* chimeric or macroconverted genes was narrowed to c.342C>T, for type 5 chimera to c.939+11G>C, and for the converted allele in sample 15 (like chimera 4) to c.292+33A>C. On the other hand, type 1 *TNXA*/*TNXB* chimeras or macroconverted genes had a start and stop points at c.11417A>G and c.11387-9T, respectively, and in *TNXA*/*TNXB* chimera type 2, at c.11616G>A and c.11548C. This analysis also allowed us to accurately define the breakpoints involved in alleles with a 3’ macroconversion and to determine that all the breakpoints were conserved for the different types of recombinant alleles.

**Figure 4:**
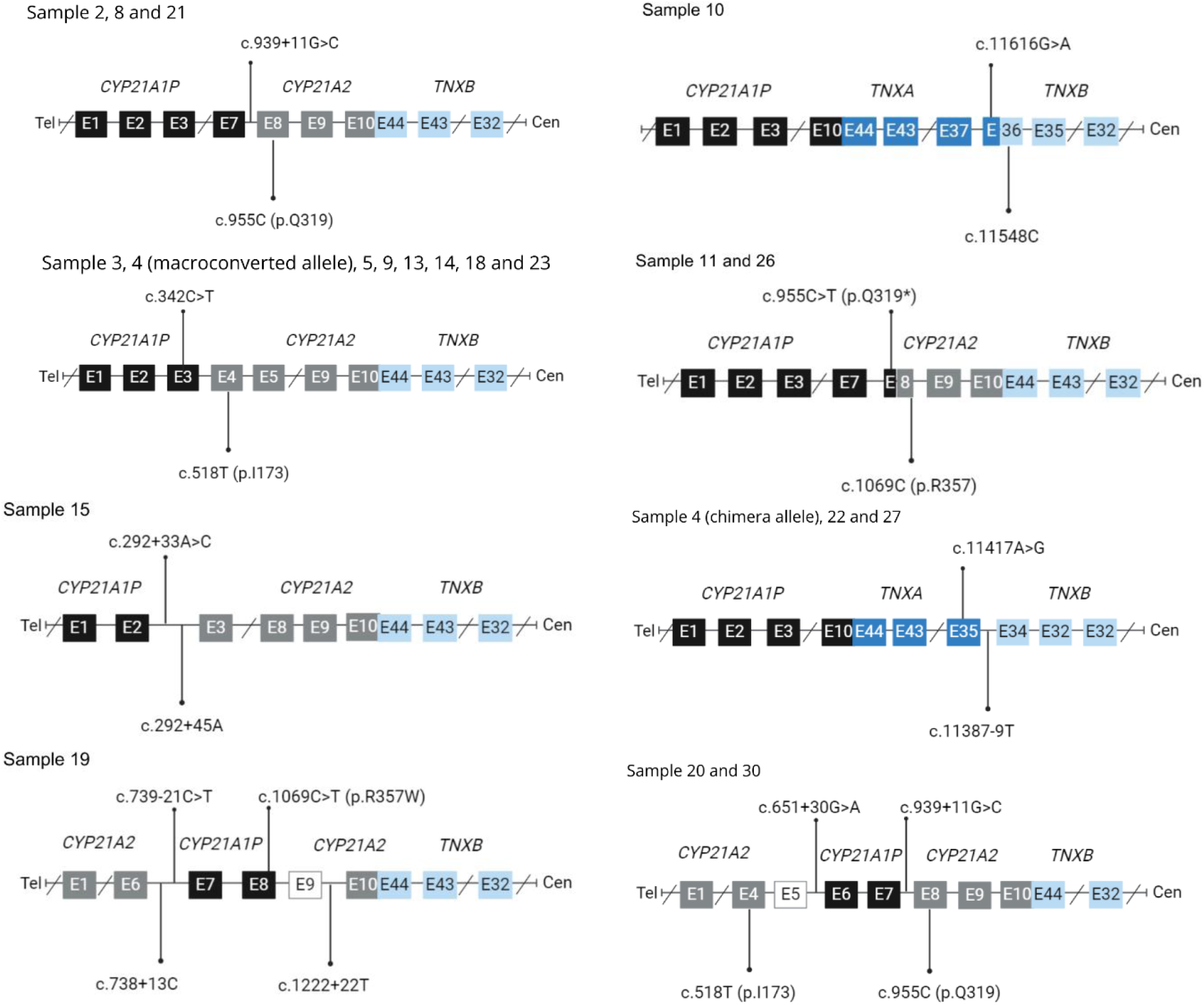
Schematic representation of the breakpoints of the different chimeras and macroconverted genes in the analyzed samples for Amplicon. **A.** *CYP21A1P* exons are shown in black boxes, *CYP21A2* exons in gray boxes, *TNXA* exons in blue boxes and *TNXB* exons in light blue boxes. White boxes represent the uncertain origin of the sequence. Above the boxes are the GVs from *CYP21A1P or TNXA* that limit the breakpoints, and below the GVs from *CYP21A2 or TNXB*. E: Exon. Tel: telomere. Cen: centromere

Of note, the initial classification of the type of chimeric or macroconverted alleles assigned, had changed for some samples following LR sequencing. One patient (sample 10) had been previously classified as a carrier of a chimera *CYP21A1P/CYP21A2* type 5, but actually the rearrangement extends up to the *TNXB* reclassifying it as a *TNXA*/*TNXB* type 2 chimera. Likewise, another patient (sample 26) has been previously genotyped as having a macroconverted allele and a chimera, both of them involving a *CYP21A1P*/*CYP21A*2 rearrangement up to p.L308Ffs*6 GV as tested by MLPA. Once again, LR sequencing allowed us to extend the breakpoints to p.Q319* and reclassified to chimera/conversion type 3. Additionally, we could uncover that all the GVs from Amplicon A in this sample are in homozygosity, despite the fact that one allele presented a macroconversion and the homologous allele a chimera. Although consanguinity is not evident for this family, we also found the p.H63L GV in both alleles and in the only one copy of the pseudogene.

We have previously observed that in many patients with p.V282L/p.V282L genotype, the entire gene showed homozygosity for common non-pathogenic variants. After analyzing LR sequencing of some samples with this genotype (samples 12, 16, 28, 31, and 33), we conclude that the entire amplicon A presented homozygosity and thus phasing and haplotyping suggested the existence of only one allele. Amplicon B, on the other hand, could be separated into 2 different haplotypes in all the analyzed samples, in accordance with the presence of at least 2 RCCX modules per chromosome. In addition, we confirm that in an RCCX module conformation with a duplicated *CYP21A2,* either wild type or having the c.293-13C>G, the p.Q319* in *CYP21A2* and the c.12469+2T>C in *TNXB* are always in the centromeric copy.

Some discordant results were apparently found in the conformation of amplicon B for some previously genotyped samples (Table 1). For example, previous MLPA results of sample 1 showed a bimodular conformation of the RCCX module for both chromosomes. However, LR output suggested that one of the alleles had a monomodular arrangement lacking the *CYP21A1P.* Note, however, that the results may also be interpreted as a bimodular arrangement and homozygosity for amplicon B. Likewise, it is well documented that p.V282L is part of a known conserved haplotype with a pseudogene duplication [32–36]. Although MLPA results confirm this rearrangement for some of the samples studied, LR showed mostly a bimodular arrangement. Nevertheless, pseudogene duplication can be inferred, in some instances, by the presence of heterozygous GVs in one of the haplotypes after phasing amplicon B.

By including the analysis by LR of amplicon B, we also analyzed the frequencies of the different GVs in *CYP21A1P* and *TNXA* (Figure 5). Of note, 19 GVs for *CYP21A1P* and 28 GVs for *TNXA* were not previously described in Latino American populations with similar ethnic origin than in our country (Supplementary Table S2). We also found 1 GV in *TNXA,* n.322-82G>C, in 11 alleles that have not been reported in any of the consulted databases. In addition, when analyzing the most frequent pseudogene variants, all the *CYP21A1P* in the analyzed samples have the c.-126T; c.-113A; c.-110C; c.-103G GVs in the promoter region, the c.293-13G in intrón 2, the p.237N-p.238E-p.240K (Cluster exon 6) and the c.923dup (Ins T in exon 7). Nevertheless, for the remaining common pseudogene GVs, we noticed a considerable degree of variability by means of their absence as part of the genomic sequences (Table 1). Besides the above mentioned allele lacking the p.G111Vfs*21 (n.331_332insGAGACTAC), 3 alleles lacks the p.L31 (n.92T>C), 3 the p.173N (n.510A>T), 15 the p.282L (n.836T>G), 13 the p.319* (n.948T>C) and 30 lacks the p.357W (n.1062T>C) GV. On the other hand, none of the *CYP21A1P* presented the p.454S (n.1353C>T), 3 alleles had the p.63L (n.188A>G), and the p.492S (n.1474A>G) variant was found present in 21 alleles. In addition, all the *TNXA* genes have the 121 bp deletion, but similarly to the results for the *CYP21A1P,* not all the alleles had the most common pathogenic variants. Indeed, 12 alleles lack the frequent c.12524A GV, 5 the c.12218A, 10 the c.12514A, and the c.12174G GV is absent in 16 alleles.

**Figure 5:**
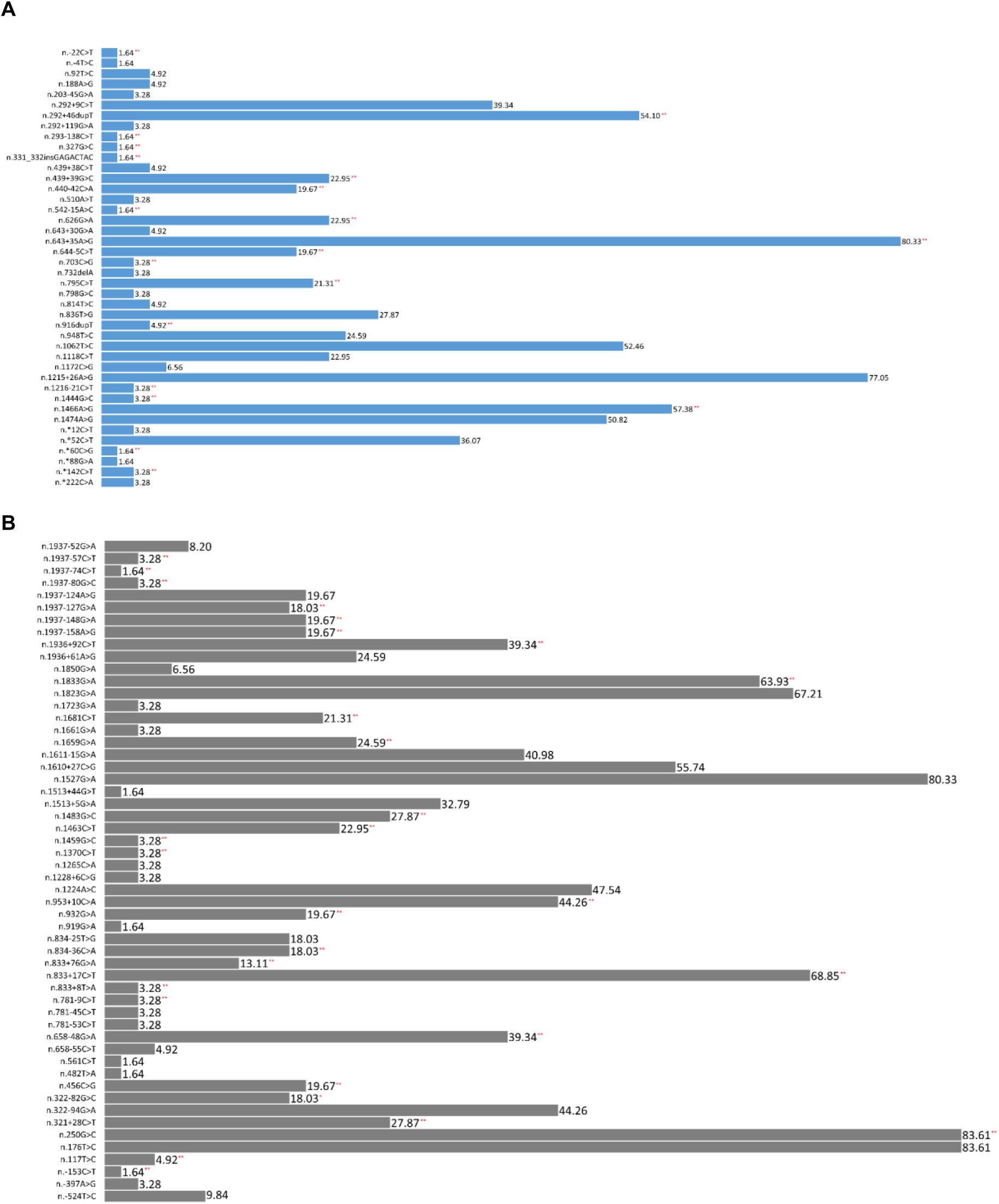
Alleles frequencies of different GVs found in amplicon B. **A:** Allele frequencies for *CYP21A1P* GVs**. B:** Allele frequencies for *TNXA* GVs. N= 61 alleles. *: SNV not reported in the consulted databases. **: GVs not reported for individuals from Latino American populations with mostly European and Native American ancestry.

Finally, we are describing a novel GV NM_000500.9:c.1208_1209del, p.H403Rfs*5 in a SV patient (sample 24). The CAT>C deletion has not been reported previously neither in population databases nor in the literature. The GV was classified as pathogenic according to the ACMG guidelines as it introduces a change in the reading frame and the appearance of a premature stop codon 5 aminoacids ahead (See Supplementary Text S1 and Supplementary Figure S2 for more details).

## DISCUSSION

In this work, we analyzed the utility of ONT third-generation sequencing for the study of GVs and structural rearrangements of the RCCX modules implicated in the CAH due to 21OHD and in CAH-X. We include samples previously studied in our laboratory and new ones, encompassing SW, SV and NC clinical forms. To our knowledge, this work represents the first study analyzing CAH patients from Latin America using LR sequencing. This includes a detailed analysis of the RCCX module containing the *CYP21A1P* and *TNXA* pseudogenes, adding valuable information of the distribution and frequencies of GVs of this genomic region in Latin American populations, barely reported in clinical and population databases.

The human RCCX module represents a complex genomic region with a high variability in the number of copies per allele. Although 2/3 of the chromosomes analyzed have a duplicated RCCX module, monomeric arrangements and up to four copies of the module have been described [37]. In addition, macro and microconversions of genomic sequences are frequently found depending on the stretch of sequences involved. In that scenario, presence of more than one pathogenic GV in *cis* is very frequent in 21OHD [12]. In conventional molecular methodologies, most of the cis/trans location of GVs requires the study of segregated parents, specially for NC patients, for an accurate genetic counseling, and in many instances they are not available. LR sequencing has the advantage that the study of segregated parents is not mandatory, as the *cis*/*trans* locations of GVs can be detected in a single read.

By using ONT LR sequencing we found concordant results for the known GVs in previously analyzed samples from our cohort. Nevertheless, data from LR allowed to complete the GVs that were not detected for some rearrangements with the current algorithm and were otherwise assumed. Moreover, we observed that common pseudogene GVs were absent in a considerable number of the rearrangements, pointing to the diversity of the sequences involved and in accordance with the variability observed when the *CYP21A1P* and *TNXA* were analyzed in our samples. In line with these observations, we observed a converted *TNXA/TNXB* allele lacking the p.G111Vfs*21 GV in a patient with a NC phenotype. This GV was also absent in the *CYP21A1P* from the same sample. Although this is a very rare haplotype (0.0024% individuals in GnomAD), the importance of this finding relies on the fact that the majority of the molecular diagnosis methods -including ours-take advantage of the absence of this 8 bp deletion to differentially amplify the *CYP21A2* gene. LR sequencing overcomes this limitation, avoiding the amplification of a pseudogene lacking this GV and a false positive result. Indeed, we have previously genotyped this sample as having p.I173N; p.I237N-p.V238E-p.M240K; p.V282L; p.L308Ffs*6; p.Q319* GVs, and thus classified it as a 3’ macroconversion. In contrast, MLPA showed a complete macroconversion up to *TNXB* with a conflicting result for the p.G111Vfs*21 GV suggesting a rare macroconverted allele. LR allowed us to verify a *TNXA/TNXB* type1 rearrangement for this sample. In addition, we conclude that the results found by Sanger sequencing may actually represent the amplification of the 3’ fragment of the converted allele, but may also include the pseudogene lacking the p.G111Vfs*21 GV.

Chimeric and large gene conversions represent a significant number of disease causing alleles in 21OHD and in CAH-X as well [37,38]. In this work, LR revealed with higher precision the breakpoints of the rearrangements adding to the (re)classification of the type of chimeric genes. In that sense, one of the most important results of our study for the clinic is the finding of a previously chimeric *CYP21A1P/CYP21A2* type 5 allele in a patient that was reclassified to a *TNXA/TNXB* type 2 chimera. These new findings change the genetic counseling for the patient and his family related to the presence of a segregated allele for CAH-X. It should be noted that only the *TNXA/TNXB* type 1 chimera can be detected using SALSA MLPA Probemix P050 CAH. Hence, patients with other *TNXA/TNXB* chimeras are missing by applying this method, triggering the search of pathogenic GVs in *TNXB* or other types of rearrangements only in the presence of clinical manifestations of CAH-X. Furthermore, for a more accurate definition of the breakpoints of the chimeric and macroconverted genes, we also took advantage of other GVs differentially described in population databases compared to the active genes. This adds to the definition made with the most frequent pathogenic GVs in the pseudogene for narrowing the sequences involved in the rearrangements.

The most frequent pathogenic GV associated with alleles in NC patients is p.V282L [1]. Thus, homozygosity for this GV is quite common in this clinical form of 21OHD, and we have previously observed that this homozygosity involved the entire *CYP21A2* gene in a considerable number of patients. LR sequencing revealed that this homozygosity extends to the neighboring *TNXB* gene in all of the analyzed samples, but not to the RCCX module containing the pseudogenes, pointing out to a conserved haplotype for the active genes. Of note, routine molecular studies in homozygous p.V282L patients may not include the analysis of GVs in the *TNXB* gene. Therefore, a detailed analysis of additional samples using LR sequencing could contribute to a more precise characterization of disease-causing alleles and to the knowledge of putative founder events.

As we have mentioned above, we found some apparent discordant results when defining the number of the RCCX modules in the analyzed samples. Most of these differences might be explained by the fact that LR sequencing of amplicons cannot eventually separate haplotypes if both copies are homozygous, and therefore, homozygosity may be indistinguishable from a putative deletion of the region. Likewise, if an identical pseudogene is duplicated in the same allele, we cannot differentiate them. Nevertheless, and although we do not phase more than 2 haplotypes for each amplicon, presence of GVs in heterozygosis in one haplotype allowed us to posit that the RCCX module containing the pseudogene is indeed duplicated in some samples.

We also aimed at analyzing in detail the GVs in the pseudogenes *CYP21A1P* and *TNXA.* As these regions are not included in the routine analysis in patients, its variability is mostly limited to the information deposited in population databases from whole exomes or genomes initiatives. In that sense, we have identified some GVs not previously described for Latin American populations in addition to a variant not reported in any of the consulted databases. Although the number of alleles sequenced may not be high enough to give a more precise estimation of its frequencies, this data may add to the characterization of the sequence diversity in different populations, and should be considered as the basis for further analysis for the study of alleles frequencies in individuals from our region.

## Conclusions

To our knowledge, this is the first work in Latin America that uses LR sequencing for analyzing a specific human disease genomic region. Using LR sequencing, we were able to confirm all the GVs previously observed by other methods, adding valuable information of the GVs involved, narrowing genetic breakpoints, and redefining genomic regions involved in rearrangements. It was also possible to determine with high confidence the phase of each allele, allowing for the first time the analysis of GVs in neighboring genes with putative clinical implications in a single read. In addition, we present data of the frequencies of GVs in the RCCX module containing the pseudogenes, contributing to the characterization of this complex genomic region.

## Supporting information

Claps_et_al_2024_Supplementary Figures

Claps_et_al_2024_Supplementary Text and Tables

## Data Availability

All data produced in the present study are available upon reasonable request to the authors

## Data Availability Statement

The original contributions presented in the study are included in the article/supplementary material, further inquiries can be directed to the corresponding author.

## Conflicts of interest

The authors declare no conflict of interest.

## Fundings

This work was supported by grants from the University of Buenos Aires (PIDAE 2020 and 2022, LD), from the National Agency for the Promotion of Science and Technology (ANPCyT, PICT-2021-CAT-II-00018, LK), and from the Administración Nacional de Laboratorios e Institutos de Salud (FOCANLIS 2022, NRU:2022-5, MT).

## Acknowledgements

We thank Dr Verónica Ferreiro for helpful discussion of the results. AC was a fellow of the Salud Investiga Programe, Ministerio de Salud de la Nación. JEK and MLI are Ph.D fellows from the National Research Council (CONICET). NM, LK and LD are researchers from the National Research Council (CONICET). AC, TC, JL, MT and LD are professional staff from the Administración Nacional de Laboratorios e Institutos de Salud (ANLIS).

## Notes

### Competing Interest Statement

The authors have declared no competing interest.

### Author Declarations

The protocol was approved and gave by the ethics committee of the Administracion Nacional de Laboratorios e Institutos de Salud (A.N.L.I.S.), Buenos Aires, Argentina

## References

1. Claahsen-van der Grinten HL, Speiser PW, Ahmed SF, Arlt W, Auchus RJ, Falhammar H, et al. Congenital Adrenal Hyperplasia-Current Insights in Pathophysiology, Diagnostics, and Management. Endocr Rev. 2022;43: 91–159.

2. White PC, New MI, Dupont B. Structure of human steroid 21-hydroxylase genes. Proc Natl Acad Sci U S A. 1986;83: 5111–5115.

3. Donohoue PA, van Dop C, McLean RH, White PC, Jospe N, Migeon CJ. Gene conversion in salt-losing congenital adrenal hyperplasia with absent complement C4B protein. J Clin Endocrinol Metab. 1986;62: 995–1002.

4. Higashi Y, Tanae A, Inoue H, Fujii-Kuriyama Y. Evidence for frequent gene conversion in the steroid 21-hydroxylase P-450(C21) gene: implications for steroid 21-hydroxylase deficiency. Am J Hum Genet. 1988;42: 17–25.

5. Chen W, Xu Z, Sullivan A, Finkielstain GP, Van Ryzin C, Merke DP, et al. Junction site analysis of chimeric CYP21A1P/CYP21A2 genes in 21-hydroxylase deficiency. Clin Chem. 2012;58: 421–430.

6. Lao Q, Burkardt DD, Kollender S, Faucz FR, Merke DP. Congenital adrenal hyperplasia due to two rare CYP21A2 variant alleles, including a novel attenuated CYP21A1P/CYP21A2 chimera. Mol Genet Genomic Med. 2023;11: e2195.

7. Adachi E, Nakagawa R, Tsuji-Hosokawa A, Gau M, Kirino S, Yogi A, et al. A MinION-based Long-Read Sequencing Application With One-Step PCR for the Genetic Diagnosis of 21-Hydroxylase Deficiency. J Clin Endocrinol Metab. 2024;109: 750–760.

8. Lee H-H, Lee Y-J, Chao M-C. Comparing the Southern blot method and polymerase chain reaction product analysis for chimeric RCCX detection in CYP21A2 deficiency. Anal Biochem. 2010;399: 293–298.

9. Carrozza C, Foca L, De Paolis E, Concolino P. Genes and Pseudogenes: Complexity of the RCCX Locus and Disease. Front Endocrinol (Lausanne). 2021;12: 709758.

10. Miller WL, Merke DP. Tenascin-X, Congenital Adrenal Hyperplasia, and the CAH-X Syndrome. Horm Res Paediatr. 2018;89: 352–361.

11. Simonetti L, Bruque CD, Fernández CS, Benavides-Mori B, Delea M, Kolomenski JE, et al. CYP21A2 mutation update: Comprehensive analysis of databases and published genetic variants. Hum Mutat. 2018;39: 5–22.

12. Fernández CS, Taboas M, Bruque CD, Benavides-Mori B, Belli S, Stivel M, et al. Genetic characterization of a large cohort of Argentine 21-hydroxylase Deficiency. Clin Endocrinol (Oxf). 2020;93: 19–27.

13. Tyson JR, O’Neil NJ, Jain M, Olsen HE, Hieter P, Snutch TP. MinION-based long-read sequencing and assembly extends the reference genome. Genome Res. 2018;28: 266– 274.

14. Karaoğlan M, Nacarkahya G, Aytaç EH, Keskin M. Challenges of CYP21A2 genotyping in children with 21-hydroxylase deficiency: determination of genotype-phenotype correlation using next generation sequencing in Southeastern Anatolia. J Endocrinol Invest. 2021;44: 2395–2405.

15. Lee H-H, Lee Y-J, Lin C-Y. PCR-based detection of the CYP21 deletion and TNXA/TNXB hybrid in the RCCX module. Genomics. 2004;83: 944–950.

16. Parajes S, Quinteiro C, Domínguez F, Loidi L. High frequency of copy number variations and sequence variants at CYP21A2 locus: implication for the genetic diagnosis of 21-hydroxylase deficiency. PLoS One. 2008;3: e2138.

17. Welcome to Oxford Nanopore Technologies. In: Oxford Nanopore Technologies [Internet]. [cited 6 Nov 2024]. Available: https://nanoporetech.com/

18. Li H. Minimap2: pairwise alignment for nucleotide sequences. Bioinformatics. 2018;34: 3094–3100.

19. Li H. Minimap and miniasm: fast mapping and de novo assembly for noisy long sequences. Bioinformatics. 2016;32: 2103–2110.

20. Releases · samtools/samtools. In: GitHub [Internet]. [cited 6 Nov 2024]. Available: https://github.com/samtools/samtools/releases

21. Zheng Z, Li S, Su J, Leung AW-S, Lam T-W, Luo R. Symphonizing pileup and full-alignment for deep learning-based long-read variant calling. Nature Computational Science. 2022;2: 797–803.

22. Martin M, Patterson M, Garg S, Fischer SO, Pisanti N, Klau GW, et al. WhatsHap: fast and accurate read-based phasing. bioRxiv. 2016. p. 085050. doi:10.1101/085050

23. den Dunnen JT, Dalgleish R, Maglott DR, Hart RK, Greenblatt MS, McGowan-Jordan J, et al. HGVS Recommendations for the Description of Sequence Variants: 2016 Update. Hum Mutat. 2016;37: 564–569.

24. Chen S, Francioli LC, Goodrich JK, Collins RL, Kanai M, Wang Q, et al. A genomic mutational constraint map using variation in 76,156 human genomes. Nature. 2023;625: 92–100.

25. Dunham I, Kulesha E, Iotchkova V, Morganella S, Birney E. FORGE: A tool to discover cell specific enrichments of GWAS associated SNPs in regulatory regions. F1000Research. 2015;4: 18.

26. Sherry ST, Ward MH, Kholodov M, Baker J, Phan L, Smigielski EM, et al. dbSNP: the NCBI database of genetic variation. Nucleic Acids Res. 2001;29: 308–311.

27. ALFA: Allele Frequency Aggregator. [cited 6 Nov 2024]. Available: www.ncbi.nlm.nih.gov/snp/docs/gsr/alfa/

28. Landrum MJ, Lee JM, Riley GR, Jang W, Rubinstein WS, Church DM, et al. ClinVar: public archive of relationships among sequence variation and human phenotype. Nucleic Acids Res. 2014;42: D980–5.

29. Rehm HL, Berg JS, Brooks LD, Bustamante CD, Evans JP, Landrum MJ, et al. ClinGen--the Clinical Genome Resource. N Engl J Med. 2015;372: 2235–2242.

30. Firth HV, Richards SM, Paul Bevan A, Clayton S, Corpas M, Rajan D, et al. DECIPHER: Database of Chromosomal Imbalance and Phenotype in Humans Using Ensembl Resources. The American Journal of Human Genetics. 2009;84: 524–533.

31. Richards S, Aziz N, Bale S, Bick D, Das S, Gastier-Foster J, et al. Standards and guidelines for the interpretation of sequence variants: a joint consensus recommendation of the American College of Medical Genetics and Genomics and the Association for Molecular Pathology. Genet Med. 2015;17: 405–424.

32. Laron Z, Pollack MS, Zamir R, Roitman A, Dickerman Z, Levine LS, et al. Late onset 21-hydroxylase deficiency and HLA in the Ashkenazi population: a new allele at the 21-hydroxylase locus. Hum Immunol. 1980;1: 55–66.

33. Pollack MS, Levine LS, O’Neill GJ, Pang S, Lorenzen F, Kohn B, et al. HLA linkage and B14, DR1, BfS haplotype association with the genes for late onset and cryptic 21-hydroxylase deficiency. Am J Hum Genet. 1981;33: 540–550.

34. Kohn B, Levine LS, Pollack MS, Pang S, Lorenzen F, Levy D, et al. Late-onset steroid 21-hydroxylase deficiency: a variant of classical congenital adrenal hyperplasia. J Clin Endocrinol Metab. 1982;55: 817–827.

35. Werkmeister JW, New MI, Dupont B, White PC. Frequent deletion and duplication of the steroid 21-hydroxylase genes. Am J Hum Genet. 1986;39: 461–469.

36. Garlepp MJ, Wilton AN, Dawkins RL, White PC. Rearrangement of 21-hydroxylase genes in disease-associated MHC supratypes. Immunogenetics. 1986;23: 100–105.

37. Chen W, Xu Z, Nishitani M, Van Ryzin C, McDonnell NB, Merke DP. Complement component 4 copy number variation and CYP21A2 genotype associations in patients with congenital adrenal hyperplasia due to 21-hydroxylase deficiency. Hum Genet. 2012;131: 1889–1894.

38. Koppens PFJ, Hoogenboezem T, Degenhart HJ. Carriership of a defective tenascin-X gene in steroid 21-hydroxylase deficiency patients: TNXB -TNXA hybrids in apparent large-scale gene conversions. Hum Mol Genet. 2002;11: 2581–2590.

39. Lindeboom RGH, Supek F, Lehner B. The rules and impact of nonsense-mediated mRNA decay in human cancers. Nat Genet. 2016;48: 1112–1118.

40. Abou Tayoun AN, Pesaran T, DiStefano MT, Oza A, Rehm HL, Biesecker LG, et al. Recommendations for interpreting the loss of function PVS1 ACMG/AMP variant criterion. Hum Mutat. 2018;39: 1517–1524.

