## Supplementary material for "HIGH PRECISION CHARACTERIZATION OF RCCX REARRANGEMENTS IN A 21-HYDROXYLASE DEFICIENCY LATIN AMERICAN COHORT USING OXFORD NANOPORE LONG READ SEQUENCING": Claps_et_al_2024_Supplementary Figures

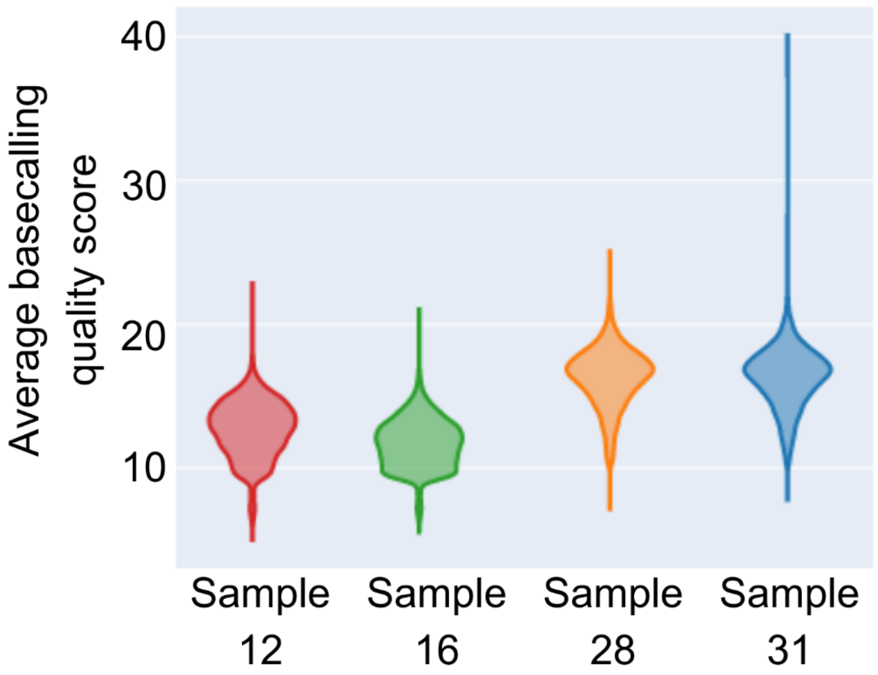


**Figure S1: Average basecalling quality score of amplicon A of four samples genotyped p.V282L/p.V282L.** Sample IDs 12 and 16 were sequenced by MinION-based long-read sequencing using Nanopore Flow Cell R9.4.1 and sample IDs 28 and 31 by PromethION-based long-read sequencing using Nanopore Flow Cell R10.4.1. The violin plots show high average basecalling quality scores in sample IDs 28 and 31.

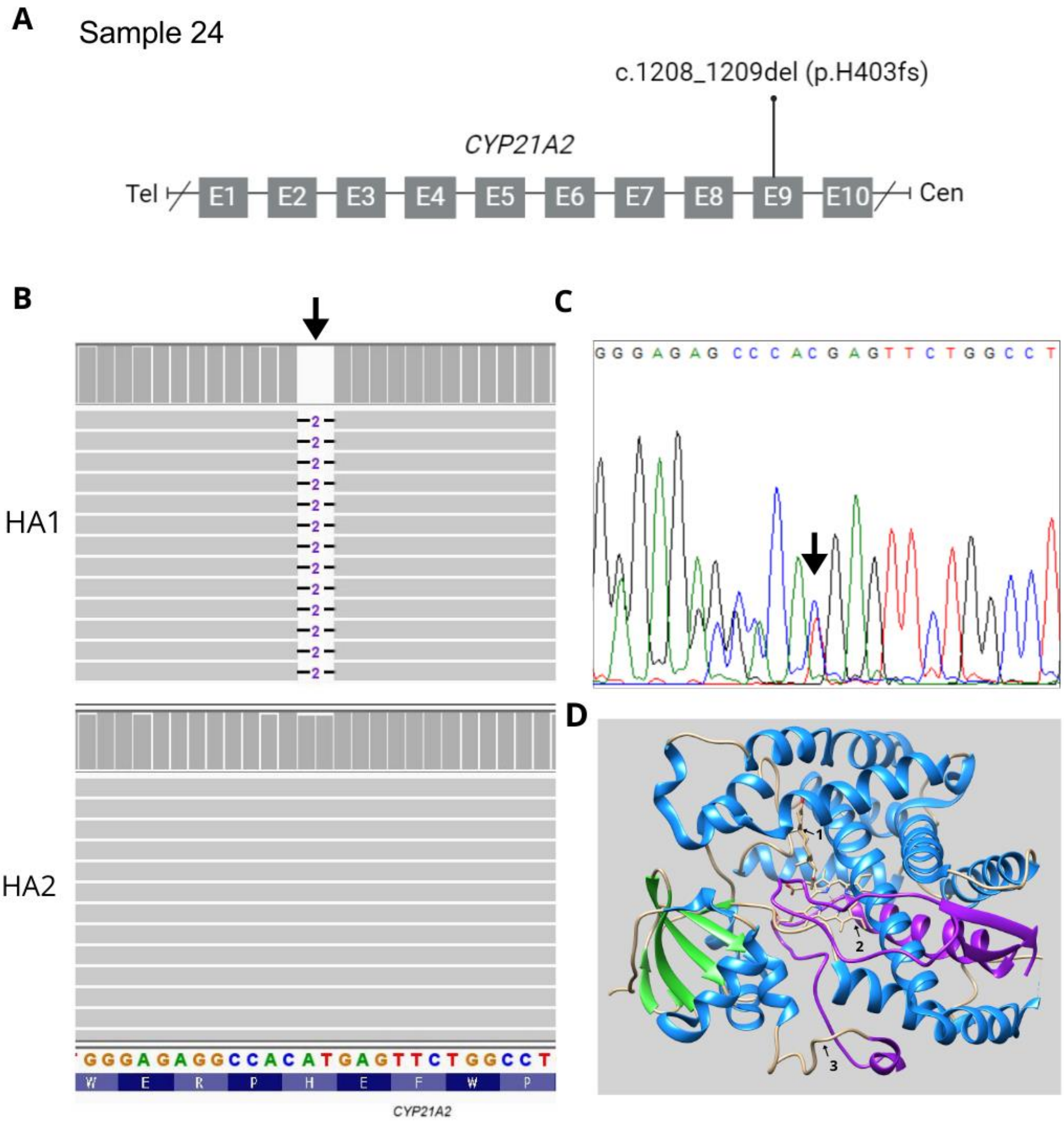


**Figure S2: Novel c.1208_1209del variant found in sample ID 24**: **A:** Schematic representation of the *CYP21A2* gene showing the location of the variant in exon 9 (arrow). **B:** Partial IGV plots of Amplicon A, HA1 (above) and HA2 (below). The arrow indicates the position of the variant in HA1. The reference nucleotide and protein sequences are indicated below. **C:** Representative partial reverse complement electropherogram of Sanger sequencing. The arrow indicates the starting point of the deletion leading to a frameshift in heterozygocis. **D:** Structure of the protein with progesterone (arrow 1) and the heme group (arrow 2) showing the p.H403 (arrow 3). In purple, the potentially 78 residues that may be absent due to the c.1208_1209del leading to a frameshift with a premature stop codon 5 aminoacids ahead. Modified from https://doi.org/10.2210/pdb4Y8W/pdb
