## Supplementary material for "HIGH PRECISION CHARACTERIZATION OF RCCX REARRANGEMENTS IN A 21-HYDROXYLASE DEFICIENCY LATIN AMERICAN COHORT USING OXFORD NANOPORE LONG READ SEQUENCING": Claps_et_al_2024_Supplementary Text and Tables

**Text S1:** The NM_000500.9:c.1208_1209del, p.H403Rfs*5 found in sample 24 is located in the exon 9 of the *CYP21A2* gene leading to changes in the residues of the protein and the appearance of a premature stop codon 5 aminoacids ahead. According to DECIPHER and ClinGen, this change may escape nonsense mediated decay (NMD) [39,40] and therefore it is expected to result in a truncated protein. By analyzing the structure of the enzyme (PDB ID 4Y8W https://doi.org/10.2210/pdb4Y8W/pdb), p.H403 is located in a loop preceding an alpha-helix that is in contact with the Heme group (Supplementary Figure S2). Absence of this critical region of the protein may lead to a null or very low enzymatic residual activity allele. Accordingly, the patient presented a SV form of the disease with ambiguous genitalia, a basal 17 hydroxyprogesterone >38 ng/mL, and the c.293-13C>G on the homologous alleles.

### Supplementary Tables

**Table S1: GVs found by LR sequencing in *CYP21A2* and *TNXB* genes in the analyzed samples**

| ***CYP21A2*** | | | |
|---|---|---|---|
| **c.** | **p.** | **c.** | **p.** |
| c.-685G>A | Promoter | c.293-13C>A | Intronic |
| c.-551A>G | Promoter | c.293-13C>G | Intronic |
| c.-536C>T | Promoter | c.293-86_293-85insGA | Intronic |
| c.-448A>G | Promoter | c.293-79G>T | Intronic |
| c.-444dup | Promoter | c.293-79G>A | Intronic |
| c.-308G>C | Promoter | c.293-74G>A | Intronic |
| c.-296T>C | Promoter | c.293-67C>A | Intronic |
| c.-295A>C | Promoter | c.293-67C>G | Intronic |
| c.-289T>C | Promoter | c.293-48A>G | Intronic |
| c.-284A>G | Promoter | c.293-44G>T | Intronic |
| c.-282T>G | Promoter | c.293-39C>G | Intronic |
| c.-210T>C | Promoter | c.293-38A>G | Intronic |
| c.-199C>T | Promoter | c.293-13C>A | Intronic |
| c.-190dup | Promoter | c.293-13C>G | Intronic |
| c.-126C>T | Promoter | c.308G>A | p.R103K |
| c.-113G>A | Promoter | c.327C>G | p.S109= |
| c.-110T>C | Promoter | c.332_339del | p.G111Vfs*21 |
| c.-103A>G | Promoter | c.342C>T | p.S114= |
| c.-4C>T | Promoter | c.447+38C>T | Intronic |
| c.-190dup | Promoter | c.518T>A | p.I173N |
| c.-126C>T | Promoter | c.550-15C>A | Intronic |
| c.-113G>A | Promoter | c.550-8T>C | Intronic |
| c.-110T>C | Promoter | c.552C>G | p.D184E |
| c.-103A>G | Promoter | c.651+30G>A | Intronic |
| c.-4C>T | Promoter | c.651+35A>G | Intronic |

| ***TNXB*** | |
|---|---|
| **c.** | **p.** |
| c.*163G>A | 3'UTR |
| c.*155G>C | 3'UTR |
| c.*127C>T | 3'UTR |
| c.12633+92T>C | Intronic |
| c.12530G>A | p.S4177N |
| c.12520G>A | p.D4174N |
| c.12469+2T>C | Intronic |
| c.12307+27C>G | Intronic |
| c.12224G>A | p.R4075H |
| c.12210+43T>G | Intronic |
| c.12210+39dup | Intronic |
| c.12210+5G>A | Intronic |
| c.12180C>G | p.C4060W |
| c.12170A>T | p.N4057I |
| c.12156C>G | p.R4052= |
| c.12058+15G>A | Intronic |
| c.12058+11G>A | Intronic |
| c.12011T>C | p.M4004T |
| c.11925+6C>G | Intronic |
| c.11921A>C | p.N3974T |
| c.11650+10C>G | Intronic |
| c.11629G>A | p.V3877I |
| c.11616G>A | p.V3872= |
| c.11605dup | p.Q3869Pfs*8 |
| c.11548C>A | p.Q3850K |

**Table S1: GVs found by LR sequencing in *CYP21A2* and *TNXB* genes in the analyzed samples**

| *CYP21A2* | | | |
|---|---|---|---|
| **c.** | **p.** | **c.** | **p.** |
| c.29_31del | p.L10del | c.705T>C | p.D235= |
| c.92C>T | p.P31L | c.710T>A | p.I237N |
| c.118C>T | p.L40= | c.711C>G | p.I237M |
| c.138C>A | p.P46= | c.713T>A | p.V238E |
| c.188A>T | p.H63L | c.719T>A | p.M240K |
| c.292+9C>T | Intronic | c.738+12A>G | Intronic |
| c. 292+34_292+36dup | Intronic | c.738+13C>T | Intronic |
| c.292+33A>C | Intronic | c.739-21C>T | Intronic |
| c. 292+45_292+46insTGT | Intronic | c.747C>G | p.L249= |
| c.292+46A>T | Intronic | c.806G>C | p.S269T |
| c.292+56T>G | Intronic | c.822T>C | p.S274= |
| c.292+67C>T | Intronic | c.844G>T | p.V282L |
| c.292+84A>G | Intronic | c.923dup | p.L308Ffs*6 |
| c.292+92A>G | Intronic | c.939+11G>C | Intronic |
| c.292+100A>G | Intronic | c.939+91G>A | Intronic |
| c.292+116A>G | Intronic | c.955C>T | p.Q319* |
| c.292+127T>G | Intronic | c.1069C>T | p.R357W |
| c. 292+137_292+138insC | Intronic | c.1119-34G>A | Intronic |
| c.292+138T>G | Intronic | c.1125C>T | p.S375= |
| c.293-139A>T | Intronic | c. 1208_1209del | p.H403Rfs*5 |

| *TNXB* | |
|---|---|
| **c.** | **p.** |
| c.11547A>G | p.T3849= |
| c.11531-25T>C | Intronic |
| c.11531-36C>A | Intronic |
| c.11531-88G>A | Intronic |
| c.11530+56G>C | Intronic |
| c.11530+46C>T | Intronic |
| c.11530+37C>T | Intronic |
| c. 11435_11524+30del | Intronic |
| c.11417A>G | p.Q3806R |
| c.11412T>C | p.D3804= |
| c.11387-9T>C | Intronic |
| c.11387-45T>C | Intronic |
| c.11264-48A>G | Intronic |
| c.11264-55T>C | Intronic |
| c.11263+39T>C | Intronic |
| c.11161C>T | p.R3721W |
| c.11142G>A | p.T3714= |
| c.11088T>A | p.T3696= |
| c.10960T>G | p.S3654A |
| c.10928-94G>A | Intronic |

**Table S1: GVs found by LR sequencing in *CYP21A2* and *TNXB* genes in the analyzed samples**

| ***CYP21A2*** | | | |
|---|---|---|---|
| **c.** | **p.** | **c.** | **p.** |
| c.293-131_293-129dup | Intronic | c.1222+22T>C | Intronic |
| c.293-123C>A | Intronic | c.1222+26G>A | Intronic |
| c.293-104dup | Intronic | c.1223-22C>T | Intronic |
| c.293-109G>C | Intronic | c.1333C>T | p.R445* |
| c.293-96_293-95del | Intronic | c.1455dup | p.M486Dfs*38 |
| c.293-93G>C | Intronic | c.1451G>C | p.R484P |
| c.293-92G>A | Intronic | c.1473G>A | p.P491= |
| c.293-86_293-85insGA | Intronic | c.1474G>A | p.G492S |
| c.293-79G>T | Intronic | c.1481G>A | p.S494N |
| c.293-79G>A | Intronic | c.*12C>T | 3'UTR |
| c.293-74G>A | Intronic | c.*13G>A | 3'UTR |
| c.293-67C>A | Intronic | c.*52C>T | 3'UTR |
| c.293-67C>G | Intronic | c.*368T>C | 3'UTR |
| c.293-48A>G | Intronic | c.*390A>G | 3'UTR |
| c.293-44G>T | Intronic | c.*440C>T | 3'UTR |
| c.293-39C>G | Intronic | c.*443T>C | 3'UTR |
| c.293-38A>G | Intronic | c.*464T>C | 3'UTR |
| | | c.*474C>T | 3'UTR |

| ***TNXB*** | |
|---|---|
| **c.** | **p.** |
| c.10927+28C>T | Intronic |
| c.10893G>A | p.K3631= |
| c.10723T>C | p.S3575P |
| c.11088T>A | p.T3696= |
| c.10677A>G | p.L3559= |
| c.10607-19C>A | Intronic |
| c.10607-256G>A | Intronic |
| c.10607-397A>G | Intronic |
| c.10606+342G>A | Intronic |
| c.10606+325C>T | Intronic |
| c.10606+262A>G | Intronic |
| c.10606+119C>G | Intronic |
| c.10606+108C>A | Intronic |
| c.10606+106C>T | Intronic |
| c.10606+61G>T | Intronic |

***CYP21A2***: RefSeq annotated: NM_000500.9|NP_000491.4 Ensembl annotated: ENST00000418967

***TNXB***: RefSeq annotated: NM_001365276.2 | NP_001352205.1. Ensembl annotated: ENST00000644971.2

**Table S2: GVs found by LR sequencing in *CYP21A1P* and *TNXA* genes in the analyzed samples and its comparison with reported LATAM frequencies**

| ***CYP21A1P*** | | | |
|---|---|---|---|
| **Genetic variant** | **Number of alleles** | **Allele frequency (%, n=61)** | **Allele frequency (%) in LATAM population[1]** |
| n.-22C>T | 1 | 1,64 | 0 |
| n.-4T>C | 1 | 1,64 | 6,9 |
| n.92T>C | 3 | 4,92 | 9 |
| n.188A>G | 3 | 4,92 | 8,9 |
| n.203-45G>A | 2 | 3,28 | 2,3 |
| n.292+9C>T | 24 | 39,34 | 58,7 |
| n.292+46dupT | 33 | 54,10 | 0 |
| n.292+119G>A | 2 | 3,28 | 14,3 |
| n.293-138C>T | 1 | 1,64 | 0 |
| n.327G>C | 1 | 1,64 | 0 |
| n.331_332insGAGACTAC | 1 | 1,64 | 0 |
| n.439+38C>T | 3 | 4,92 | 1,8 |
| n.439+39G>C | 14 | 22,95 | 0 |
| n.440-42C>A | 12 | 19,67 | 0 |
| n.510A>T | 2 | 3,28 | 3,1 |
| n.542-15A>C | 1 | 1,64 | 0 |
| n.626G>A | 14 | 22,95 | 0 |
| n.643+30G>A | 3 | 4,92 | 6 |
| n.643+35A>G | 49 | 80,33 | 0 |
| n.644-5C>T | 12 | 19,67 | 0 |
| n.703C>G | 2 | 3,28 | 0 |
| n.732delA | 2 | 3,28 | 2,1 |
| n.795C>T | 13 | 21,31 | 0 |
| n.798G>C | 2 | 3,28 | 13,9 |
| n.814T>C | 3 | 4,92 | 4,3 |
| n.836T>G | 17 | 27,87 | 23,1 |

| ***TNXA*** | | | |
|---|---|---|---|
| **Genetic variant** | **Number of alleles** | **Allele frequency (%, n=61)** | **Allele frequency (%) in LATAM population[1]** |
| n.1937-148G>A | 12 | 19,7 | 0 |
| n.1937-158A>G | 12 | 19,7 | 0 |
| n.1936+92C>T | 24 | 39,3 | 0 |
| n.1936+61A>G | 15 | 24,6 | 4,9 |
| n.1850G>A | 4 | 6,6 | 16,2 |
| n.1833G>A | 39 | 63,9 | 0 |
| n.1823G>A | 41 | 67,2 | 68 |
| n.1823G>A | 2 | 3,3 | 2 |
| n.1681C>T | 13 | 21,3 | 0 |
| n.1661G>A | 2 | 3,3 | 2 |
| n.1659G>A | 15 | 24,6 | 0 |
| n.1611-15G>A | 25 | 41,0 | 8 |
| n.1610+27C>G | 34 | 55,7 | 38,2 |
| n.1527G>A | 49 | 80,3 | 84,3 |
| n.1513+44G>T | 1 | 1,6 | 0,03 |
| n.1513+5G>A | 20 | 32,8 | 16,7 |
| n.1483G>C | 17 | 27,9 | 0 |
| n.1463C>T | 14 | 23,0 | 0 |
| n.1459G>C | 2 | 3,3 | 0 |
| n.1370C>T | 2 | 3,3 | 0 |
| n.1265C>A | 2 | 3,3 | 8 |
| n.1228+6C>G | 2 | 3,3 | 0,8 |
| n.1224A>C | 29 | 47,5 | 66,1 |
| n.953+10C>G | 27 | 44,3 | 0 |
| n.932G>A | 12 | 19,7 | 0 |
| n.919G>A | 1 | 1,6 | 2,1 |

**Table S2: GVs found by LR sequencing in *CYP21A1P* and *TNXA* genes in the analyzed samples and its comparison with reported LATAM frequencies**

| *CYP21A1P* | | | |
|---|---|---|---|
| Genetic variant | Number of alleles | Allele frequency (%, n=61) | Allele frequency (%) in LATAM population[1] |
| n.916dupT | 3 | 4,92 | 0 |
| n.948T>C | 15 | 24,59 | 7,9 |
| n.1062T>C | 32 | 52,46 | 26,4 |
| n.1118C>T | 14 | 22,95 | 6,9 |
| n.1172C>G | 4 | 6,56 | 2,5 |
| n.1215+26A>G | 47 | 77,05 | 84,8 |
| n.1216-21C>T | 2 | 3,28 | 0 |
| n.1444G>C | 2 | 3,28 | 0 |
| n.1466A>G | 35 | 57,38 | 0 |
| n.1474A>G | 31 | 50,82 | 11,6 |
| n.*12C>T | 2 | 3,28 | 0,2 |
| n.*52C>T | 22 | 36,07 | 29,3 |
| n.*60C>G | 1 | 1,64 | 0 |
| n.*88G>A | 1 | 1,64 | 1,1 |
| n.*142C>T | 2 | 3,28 | 0 |
| n.*222C>A | 2 | 3,28 | 1,5 |
| n.1937-52G>A | 5 | 8,2 | 0,2 |
| n.1937-57C>T | 2 | 3,3 | 0 |
| n.1937-74C>T | 1 | 1,6 | 0 |
| n.1937-80G>C | 2 | 3,3 | 0 |
| n.1937-124A>G | 12 | 19,7 | 0,02 |
| n.1937-127G>A | 11 | 18,0 | 0 |

| *TNXA* | | | |
|---|---|---|---|
| Genetic variant | Number of alleles | Allele frequency (%, n=61) | Allele frequency (%) in LATAM population[1] |
| n.834-25T>G | 11 | 18,0 | 7 |
| n.834-36C>A | 11 | 18,0 | 0 |
| n.833+76G>A | 8 | 13,1 | 0 |
| n.833+17C>T | 42 | 68,9 | 0 |
| n.833+8T>A | 2 | 3,3 | 0 |
| n.781-9C>T | 2 | 3,3 | 0 |
| n.781-45C>T | 2 | 3,3 | 4,1 |
| n.781-53C>T | 2 | 3,3 | 1,6 |
| n.658-48G>A | 24 | 39,3 | 0 |
| n.658-55C>T | 3 | 4,9 | 3,6 |
| n.561C>T | 1 | 1,6 | 0,7 |
| n.482T>A | 1 | 1,6 | 14,9 |
| n.456C>G | 12 | 19,7 | 0 |
| **n.322-82G>C** | 11 | 18,0 | - |
| n.322-94G>A | 27 | 44,3 | 1,5 |
| n.321+28C>T | 17 | 27,9 | 0 |
| n.250G>C | 51 | 83,6 | 0 |
| n.176T>C | 51 | 83,6 | 0,8 |
| n.117T>C | 3 | 4,9 | 0 |
| n.-153C>T | 1 | 1,6 | 0 |
| n.-397A>G | 2 | 3,3 | 1,5 |
| n.-524T>C | 6 | 9,8 | 16,9 |

1: Refers to Latin American (LATAM) individuals with mostly European and Native American ancestry (n= 610) alleles, www.ncbi.nlm.nih.gov/snp/docs/gsr/alfa/). In bold, the *TNXA* GV is not reported in any of the consulted databases.

***CYP21A1P***: Ensembl annotated: ENST00000354927. TNXA: Ensembl annotated: ENST00000507684.1
